# Menstrual Cycle Exacerbation of ADHD Symptoms in Adults: Common, Person-Specific, and Not Confined to the Luteal Phase

**DOI:** 10.64898/2026.11.09.26367690

**Authors:** Tory A. Eisenlohr-Moul, Ashley Ross, Jessica R. Peters, Anisha Nagpal, Katja Schmalenberger, Kelsey K. Wiggs, Emily Knapp, Anna Patterson, Michelle Martel

## Abstract

**Objective:** Patients often describe menstrual-cycle worsening of attention-deficit/hyperactivity disorder (ADHD). We prospectively evaluated sample- and person-level cycle effects on ADHD symptoms and their independence from cyclical negative affect and pain.

**Methods:** In an intensive longitudinal cohort (2019-2024), 106 naturally cycling adults meeting DSM-5 ADHD criteria on integrated assessment rated daily inattention and hyperactivity-impulsivity (Barkley Adult ADHD Rating Scale-IV), negative affect, and pain (median, 41 days) and provided daily salivary estradiol and progesterone for ovulation coding and phase-aligned cycle time (PACTS). Generalized additive models estimated sample-level and person-specific cycle curves; robust cyclicity was tested against each participant’s own time-shuffle null. Lab cognitive testing at four counterbalanced phases provided post hoc validation.

**Results:** Sample-average cycle effects were significant yet small (<1% of daily variance), but 54% of 87 participants tested individually were robustly cyclical in ADHD symptoms (95% CI, 44%-64%; inattention 43%, hyperactivity-impulsivity 26%), more often with informant-confirmed ADHD (69% vs 36%). Among them, the cycle explained median 52% and 51% of symptom variance, a mean swing of 2.6 and 2.1 DSM-5 criteria. Most remained cyclical after adjustment for affect (68%) or pain (81%). Worsening most often peaked at and after menses rather than premenstrually; no cycle met prospective criteria for premenstrual dysphoric disorder (PMDD). Hormones did not differ by cyclicity.

**Conclusions:** About half of these adults with ADHD showed robust cyclical symptom change, mostly not reducible to cyclical affect or pain and usually outside the premenstrual window. Future work should test its stability and whether a DSM-5 cyclical specifier is warranted.

## Introduction

Attention-deficit/hyperactivity disorder (ADHD) is a serious and debilitating neurodevelopmental disorder carrying functional, emotional, and reproductive risk (1). Clinical reports in female patients with ADHD often describe menstrual cycle–related worsening of inattention, hyperactivity-impulsivity, and emotion dysregulation (2, 3). However, such reports require prospective confirmation in daily symptom ratings, given the high false-positive rate of interview-based diagnoses in other cyclical disorders, including premenstrual dysphoric disorder (PMDD) (4, 5). If cyclical exacerbation of ADHD is prevalent, it could have important implications for ADHD conceptualizations, diagnostic methods, and treatment needs.

A further challenge is that cyclical pain and affect can mimic cyclical ADHD. Although most menstrual pain is mild and does not impair functioning, roughly one-third experience moderate-to-severe pain, often due to gynecologic disorders such as endometriosis or adenomyosis, that impairs concentration (6, 7). Cyclical affective symptoms vary similarly across individuals (8). On the severe end of the spectrum, 5.5% demonstrate premenstrual affective symptoms consistent with DSM-5 PMDD (9), whose criteria include difficulty concentrating (10), and premenstrual exacerbation occurs in up to 58% of patients with depressive disorders (8, 11). Because ADHD is highly comorbid with affective disorders, especially in female patients (12), apparent cyclical ADHD worsening could reflect cyclical pain or affect rather than ADHD itself.

The monthly menstrual cycle is characterized by recurrent fluctuations in estradiol (E2) and progesterone (P4), with pleiotropic effects across dopaminergic, noradrenergic, serotonergic, neuroimmune, and neurotrophic systems, making hormonal effects on any neuropsychiatric disorder highly plausible (13, 14). Individuals differ in neurobehavioral sensitivity to normal hormone change (15, 16). The Dimensional Affective Sensitivity to Hormones across the Menstrual Cycle (DASH-MC) framework (17) posits 3 transdiagnostic sensitivities timed to specific hormone triggers, each with a distinct prediction with respect to ADHD. <u>Perimenstrual sensitivity to low or falling E2</u> is expected to impair executive function and worsen inattention. Our prior work supports this prediction: a crossover randomized controlled trial of perimenstrual E2 stabilization found that low and falling E2 around menses (under placebo) impaired working memory in transdiagnostic psychiatric outpatients, reversible with E2 administration (18), and in a daily-diary study of a high-impulsivity community sample, E2 correlated negatively with daily ADHD symptoms (19). <u>Ovulatory sensitivity to the surge in E2</u> is expected to promote impulsive reward-seeking and worsen hyperactivity-impulsivity. <u>Luteal sensitivity to the post-ovulatory rise in P4 metabolites</u> produces primarily affective symptoms, as in PMDD, so ADHD worsening in this window may be difficult to distinguish from cyclical irritability or depression. Yet most clinical accounts of cyclical ADHD, like those of PMDD, have focused primarily on the premenstrual weeks (20, 21), rather than the menstrual and ovulatory windows where DASH-MC predicts worsening of executive function and impulsivity.

One prospective study of stimulant-treated adults found ADHD symptoms peaked during menstruation (22), but no study has estimated how many patients show clinically meaningful person-level cyclicity, when it occurs, or whether it is separable from day-to-day noise, cyclical affect, pain, or PMDD. Because retrospective reports of cyclical change are unreliable (4, 5), prospective studies are critical. We tested (1) whether, among patients with ADHD, self-reported symptoms or objective cognitive performance (inhibition, working memory) change across the cycle; (2) how much individuals differ, and whether differences correspond to specific windows or hormones; and (3) whether cyclical ADHD change is distinguishable from changes in affect or pain. We expected small sample-level changes but larger, clinically meaningful cyclic change in a subset of patients, aligned with dimensional sensitivity to varied hormonal triggers, and that cyclical ADHD change would persist after adjustment for same-day affect and pain.

## Methods

### Design and participants

This intensive longitudinal cohort study (approved by the University of Kentucky Institutional Review Board; written informed consent) was conducted 2019-2024 at the University of Kentucky (Lexington). Participants provided evening symptom ratings (ADHD, affect, pain) and morning saliva (E2, P4, and luteinizing hormone) at home for up to about 70 days (unselected start phase; about 2 cycles). Cohort size was fixed by the parent grant; power was quantified via simulation. Eligible participants were 18-35 years, naturally cycling monthly, not pregnant, breastfeeding, or using hormones, and endorsed clinically significant ADHD symptoms on the Barkley Adult ADHD Rating Scale (BAARS-IV). Participants self-reported race and ethnicity (sample descriptives). Diagnostic characterization combined the Structured Clinical Interview for DSM-5 (SCID-5), self- and informant-report on the BAARS-IV (23), and daily BAARS-IV ratings (eMethods 1). The analytic sample comprised participants meeting DSM-5 ADHD criteria (≥5 symptoms in a domain) (eFigure 1; eTables 1 and 2).

### Procedures and exposures

Ovulation was double-coded from menses onset dates and person-standardized hormone trajectories, defining ovulation as the day before a detectable P4 rise over follicular baseline (24). Using menses and ovulation as anchors, cycle time was computed using phase-aligned cycle time scaling (PACTS) (25), a method validated to align cyclical hormonal trajectories across people and cycles. Unobserved ovulations, missing closing onsets, and (in population models) onsets following each participant’s last observed onset were imputed by fixed rules (eMethods 1).

### Outcomes

Primary outcomes were daily inattention and hyperactivity-impulsivity severity (BAARS-IV item means, 0–3; within-person ω, 0.84 and 0.81; eTable 3). Secondary outcomes, negative affect (11-item Daily Record of Severity of Problems [DRSP] composite; ω, 0.91) (26) and joint/muscle pain (DRSP item), served as secondary outcomes and covariates. A daily DSM-5 count of symptoms endorsed at a clinically significant level (“often” or “very often,” 0–9) was reported as an effect size. Working memory (n-back, symmetry span, letter-number sequencing) and inhibition (Stroop, antisaccade) were measured at 4 counterbalanced in-lab visits (mid-follicular, peri-ovulatory, mid-luteal, and premenstrual phases); each visit was analyzed at its actual PACTS position (tasks and references, eMethods 6).

### Analysis

Analyses used R 4.5 with per-outcome distributional families; we report both uncorrected P and Benjamini-Hochberg (27) q values (eMethods 2). Person-level analyses used only days anchored by observed menses onset (≥10 days, ≥1 per cycle quarter; 1 exception, eMethods 1); population models used every day with PACTS cycle time. Population effects were estimated with generalized additive mixed models on PACTS cycle time, with a person-level random cyclic factor-smooth (eMethods 2). Person-specific generalized additive models (cyclic cubic spline) estimated each patient’s cycle curve; cyclic R² was defined as the share of a participant’s day-to-day variance explained by the cycle, and amplitude was the curve’s peak-to-trough difference. A participant was classified as robustly cyclical if cyclic R² or amplitude exceeded the 95th percentile of a within-person permutation null distribution (5,000 time shuffles of the participant’s own days; eMethods 3), with P values computed per Phipson and Smyth (28). The simulated false-positive rate of the cyclic R² criterion was 4.7%; power to detect a true cyclic R² of 0.3 was 59% overall and 91% with at least 30 rated days, so prevalence estimates are likely conservative (eMethods 3). Smoothing mixture models identified latent curve-shape classes matching the DASH-MC hypothesized risk windows (eMethods 4). Phase-elevation scores were calculated as the mean elevation of a participant’s own curve, relative to their cycle mean and in units of person-SD, within 3 a priori DASH-MC hormone-timed windows over PACTS time (luteal; menstrual, from menses to early follicular phase; peri-ovulatory). Mixed models related each outcome to daily E2 and P4 levels and the interaction (eMethods 5; eTable 4). Working-memory models are described in eMethods 6. Cycle-level PMDD diagnosis used the standardized Carolina Premenstrual Assessment Scoring System (4) (C-PASS; eMethods 7). Medication was examined as a moderator of cyclic change (eMethods 8).

## Results

### Sample

Of 125 participants providing daily ratings, 10 were excluded (9 under the symptom rules, 1 with too few PACTS anchors; eFigure 1; eMethods 1). Of the remaining 115, 106 had outcome days with cycle time (analytic sample), and 87 of these had adequate cycle coverage of their daily ADHD ratings for person-level analyses (Table 1; 2,494 rated days); 79 of the 87 contributed hormones (eFigure 1; eTable 1). Median daily-survey completion was 67% (interquartile range, 52%-83%); nonresponse peaked around menses but did not follow higher-symptom days (eMethods 9).

**Figure 1.**
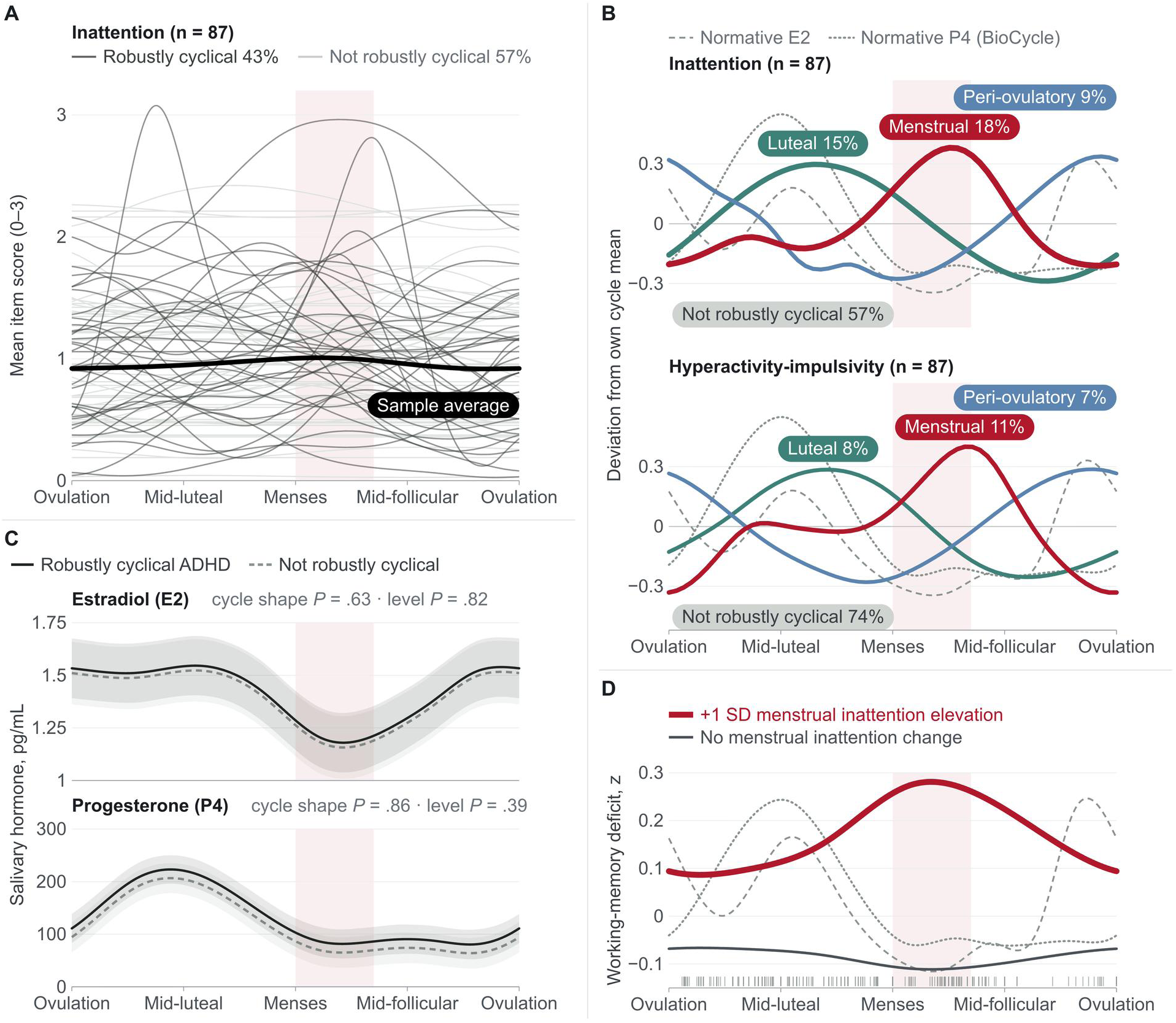
Timing of Menstrual Cycle Worsening in Adults With ADHD Symptoms, Hormone Cycles by Cyclicity Status, and Working Memory. (A) Person-specific model-implied daily inattention trajectories (N = 87; thin lines; dark = robustly cyclical [43%], light = not [57%]) around the near-flat sample average (black; population model, 106 participants); hyperactivity-impulsivity (not shown) was similar. Person-specific curves are not bounded by the 0-3 scale; one exceeds 3. (B) Where robustly cyclical participants’ own curves peaked: each was assigned the window (luteal, menstrual, or peri-ovulatory) with their highest phase-elevation score; curves are the mean deviation from own cycle mean (item-score units) of the participants peaking in each window, labels give each window’s share of all 87 participants (the remainder were not robustly cyclical; no curve shown), and line thickness increases with that share. Normative serum E2 (dashed) and P4 (dotted) from the BioCycle Study (40), modeled on the same cycle axis and rescaled to the panel, are overlaid for timing reference (also in D). (C) Model-fitted salivary E2 and P4 across the cycle in participants with (solid) and without (dashed) robust ADHD cyclicity (cyclic mixed models on the raw hormone values with a person random intercept, adjusted for age and BMI; bands = 95% CI); neither cycle shape nor level differed detectably (P values in the panels). (D) Exploratory: model-implied working-memory deficit (composite z score; higher = worse) across the cycle for a participant 1 SD above the sample mean on menstrual inattention elevation (red) and for one with no menstrual inattention change (elevation score of 0; gray; 85 participants, 321 visits). Curves are from a cyclic generalized additive mixed model in which the shape of the working-memory cycle could vary with the standardized elevation score (computed on a menstrual window taken from the inattention mixture classes [eMethods 4]; P = .06). A model constraining working memory to the shape of the normative E2 curve gave a stronger test (elevation × normative E2, P = .001; eMethods 6). Tick marks give visit positions. In all panels, cycle time is standardized (PACTS; menses onset = 0, ovulation = ±1) and the shaded band marks menses. N = 87 naturally cycling adults (A, B); 74 of the 87 with cycle-timed hormone samples and recorded age and BMI (C); 85 with in-lab visits and a per-person inattention curve (D). ADHD indicates attention-deficit/hyperactivity disorder; BMI, body mass index; E2, estradiol; P4, progesterone; PACTS, phase-aligned cycle time scaling.

**Table 1.** Participant Characteristics by Robust ADHD-Symptom Cyclicity.

| **Characteristic** | **Total (N=87)** | **Robustly cyclical ADHD (n=47)** | **Not robustly cyclical (n=40)** | **Test statistic; P value** |
|---|---|---|---|---|
| **Demographic** |  |  |  |  |
| Age, mean (SD), y | 21.5 (2.2) | 21.5 (2.4) | 21.4 (1.9) | t(80.9) = −0.21; .83 |
| Missing, No. | 4 | 2 | 2 |  |
| Sex, No. (%) |  |  |  |  |
| Female | 87 (100) | 47 (100) | 40 (100) |  |
| Race and ethnicity, No. (%) |  |  |  | χ²(4) = 6.97; .14 |
| Asian | 2 (2) | 0 (0) | 2 (5) |  |
| Black or African American | 9 (11) | 4 (9) | 5 (13) |  |
| Hispanic or Latino | 5 (6) | 5 (11) | 0 (0) |  |
| Multiracial or other | 4 (5) | 2 (4) | 2 (5) |  |
| White | 63 (76) | 34 (76) | 29 (76) |  |
| Missing | 4 |  |  |  |
| Educational attainment, No. (%) |  |  |  | χ²(3) = 3.03; .39 |
| Graduated 4-year college | 12 (15) | 5 (13) | 7 (18) |  |
| Graduated high school/high school equivalent | 2 (3) | 0 (0) | 2 (5) |  |
| Some college/trade school | 48 (62) | 27 (68) | 21 (55) |  |
| Some or completed graduate/professional school | 16 (21) | 8 (20) | 8 (21) |  |
| Missing | 9 |  |  |  |
| Marital status, No. (%) |  |  |  | χ²(2) = 4.92; .09 |
| Never married | 66 (85) | 37 (93) | 29 (76) |  |
| Married/partnered | 9 (12) | 3 (8) | 6 (16) |  |
| Previously married | 3 (4) | 0 (0) | 3 (8) |  |
| Missing | 9 |  |  |  |
| Body mass index, mean (SD) | 26.2 (7.0) | 26.3 (6.4) | 26.1 (7.6) | t(72.7) = −0.15; .88 |
| Missing, No. | 5 | 3 | 2 |  |
| Body mass index, range | 15.3-60.6 | 15.3-40.9 | 18.1-60.6 |  |
| **ADHD diagnostic characterization** |  |  |  |  |
| Met informant-confirmed ADHD criteria, No. (%) | 42 (52) | 29 (67) | 13 (34) | OR 3.91 (1.44-11.17); .004 |
| ADHD presentation (SCID), No. (%) |  |  |  | χ²(2) = 0.22; .90 |
| Combined | 54 (72) | 28 (70) | 26 (74) |  |
| Hyperactive-impulsive | 2 (3) | 1 (3) | 1 (3) |  |
| Inattentive | 19 (25) | 11 (28) | 8 (23) |  |
| Missing | 12 |  |  |  |
| SCID inattention symptom count, mean (SD) | 7.4 (1.8) | 7.5 (1.7) | 7.3 (2.0) | t(77.4) = −0.31; .76 |
| Missing, No. | 3 | 3 | 0 |  |
| SCID hyperactivity-impulsivity count, mean (SD) | 5.4 (2.1) | 5.5 (2.2) | 5.3 (2.2) | t(81.3) = −0.22; .83 |
| Missing, No. | 3 | 3 | 0 |  |
| SCID total symptom count, mean (SD) | 12.8 (3.4) | 12.9 (3.4) | 12.7 (3.5) | t(80.7) = −0.31; .76 |
| Missing, No. | 3 | 3 | 0 |  |
| **Psychiatric comorbidity (SCID-5), No. (%)** |  |  |  |  |
| Provisional SCID PMDD<sup>a</sup> | 39 (50) | 22 (55) | 17 (45) | OR 1.50 (0.56-4.05); .50 |
| Any depressive disorder | 17 (22) | 7 (18) | 10 (26) | OR 0.60 (0.17-2.01); .42 |
| Any anxiety disorder | 41 (53) | 21 (53) | 20 (53) | OR 0.99 (0.37-2.65); >.99 |
| Generalized anxiety disorder | 21 (27) | 13 (33) | 8 (21) | OR 1.79 (0.58-5.82); .31 |
| Social anxiety disorder | 21 (27) | 13 (33) | 8 (21) | OR 1.79 (0.58-5.82); .31 |
| Other anxiety disorder<sup>b</sup> | 9 (12) | 3 (8) | 6 (16) | OR 0.44 (0.07-2.25); .30 |
| Posttraumatic stress disorder | 23 (29) | 13 (33) | 10 (26) | OR 1.34 (0.45-4.06); .62 |
| Substance use disorder | 13 (17) | 7 (18) | 6 (16) | OR 1.13 (0.29-4.55); >.99 |
| **Psychotropic medication, No. (%)** |  |  |  |  |
| Stimulant | 16 (20) | 10 (24) | 6 (16) | OR 1.66 (0.48-6.25); .41 |
| Antidepressant (any) | 21 (27) | 14 (35) | 7 (18) | OR 2.36 (0.75-8.01); .13 |
| SSRI | 15 (19) | 11 (28) | 4 (11) | OR 3.18 (0.83-15.18); .08 |
| Anxiolytic | 4 (5) | 1 (3) | 3 (8) | OR 0.30 (0.01-3.98); .35 |
| Mood stabilizer | 2 (3) | 1 (3) | 1 (3) | OR 0.95 (0.01-76.42); >.99 |
| **Intensive-longitudinal data** |  |  |  |  |
| Daily-survey completion, median (IQR), % | 66.7 (51.9-83.1) | 66.7 (53.1-84.1) | 66.6 (50.0-80.3) | W = 874; .58 |
| Cycle-positioned days, median (IQR) | 40.0 (34.0-51.5) | 38.0 (33.5-50.0) | 42.5 (35.0-53.3) | W = 1,031; .44 |
| Days with hormone sample, median (IQR) | 30.0 (25.0-35.0) | 30.0 (23.5-34.0) | 32.5 (25.8-36.3) | W = 1,083; .22 |
| **Cyclicity diagnostics** |  |  |  |  |
| Robustly cyclical: inattention, No. (%) | 37 (43) | 37 (79) | 0 (0) | NA<sup>c</sup> |
| Robustly cyclical: hyperactivity-impulsivity | 23 (26) | 23 (49) | 0 (0) | NA<sup>c</sup> |
| Robustly cyclical: negative affect | 29 (33) | 24 (51) | 5 (13) | OR 7.13 (2.24-27.46); <.001 |
| Robustly cyclical: joint/muscle pain | 29 (33) | 18 (38) | 11 (28) | OR 1.63 (0.60-4.54); .36 |
| Cyclic R², negative affect, median (IQR), % | 15.4 (0.0-35.5) | 30.5 (3.0-44.4) | 2.9 (0.0-18.1) | W = 536; <.001 |
| Premenstrual elevation, maximum, median (IQR), % | 17.9 (7.9-25.7) | 15.9 (6.8-25.4) | 18.8 (8.3-29.2) | W = 800; .45 |
| Missing, No. | 10 | 3 | 7 |  |
| Premenstrual elevation ≥30%, No. (%) | 15 (19) | 7 (16) | 8 (24) | OR 0.60 (0.16-2.15); .40 |

Abbreviations: ADHD, attention-deficit/hyperactivity disorder; DSM-5, *Diagnostic and Statistical Manual of Mental Disorders*, Fifth Edition; IQR, interquartile range; OR, odds ratio; PMDD, premenstrual dysphoric disorder; SCID, Structured Clinical Interview for DSM-5; SSRI, selective serotonin reuptake inhibitor.
Participants in the person-level analyses (87 of the 106 in the analytic sample). The analytic sample of 106 had a mean (SD) age of 21.5 (2.2) years and a median 41-day diary window; all were female; all met DSM-5 ADHD criteria on integrated SCID-5, informant, and daily-report assessment, and 50 of the 93 with informant data (54%) had informant-confirmed ADHD. Robustly cyclical: inattention or hyperactivity-impulsivity beyond the participant’s own within-person permutation null on cyclic R² or amplitude (n = 47); not robustly cyclical, n = 40. Percentages use non-missing denominators (informant-confirmed ADHD, n = 81; stimulant use, n = 80; SCID-5 comorbidity and other medication rows, n = 78); pain cyclicity uses all 87 (the 4 who never reported pain are non-cyclical by definition). Test statistics: Welch t, χ², Fisher odds ratio (95% CI), or Wilcoxon W. Informant-confirmed percentages are within cyclicity groups (67% vs 34%); the text gives them within confirmation groups (69% vs 36%).
<sup>a</sup> Retrospective SCID-5 PMDD criteria endorsed at intake; not prospectively confirmed (see Results).
<sup>b</sup> Any SCID-5 anxiety disorder other than generalized or social anxiety disorder (panic disorder, agoraphobia, specific phobia); generalized and social anxiety disorder overlap, so 'any anxiety disorder' is fewer than their sum.
<sup>c</sup> Not tested: these rows define the groups (NA, not applicable).

### Sample-average cycle effects

In the analytic sample (n = 106), PACTS cycle time was associated with slight menstrual elevations of all 4 outcomes (all P and q < .001; Table 2). All population effects were small (<1% of day-to-day variance; trough-to-peak 12% to 18% of the outcome mean).

**Table 2.** Per-Outcome Cyclicity Summary.

| Outcome group | Outcome | Participants, No. | Daily reports, No. | Cycle effect, edf | P value | P value (BH) | Variance explained by cycle, % | Robustly cyclical, % (No./total); FDR survivors, No. | Cyclic R², %: mean (all); median (cyclical) | Peak window, % of robustly cyclical participants | Cyclic R² by informant-confirmed ADHD, d (P value) |
|---|---|---|---|---|---|---|---|---|---|---|---|
| Primary (ADHD) | Inattention severity | 106 | 2,580 | 3.13 | <.001 | <.001 | 0.22 | 43% (37/87); 19 | 26; 52 | Menstrual 43; luteal 35; peri-ovulatory 22 | +0.54 (.02) |
| Primary (ADHD) | Hyperactivity-impulsivity severity | 106 | 2,580 | 3.68 | <.001 | <.001 | 0.27 | 26% (23/87); 4 | 21; 51 | Menstrual 43; luteal 30; peri-ovulatory 26 | +0.41 (.07) |
| Secondary | Negative affect | 106 | 2,583 | 4.46 | <.001 | <.001 | 0.99 | 33% (29/87); 7 | 21; 45 | Menstrual 45; luteal 34; peri-ovulatory 21 | +0.34 (.13) |
| Secondary | Joint/muscle pain | 106 | 2,571 | 4.28 | <.001 | <.001 | 0.89 | 33% (29/87); 19 | 24; 57 | Menstrual 52; luteal 28; peri-ovulatory 21 | +0.41 (.07) |

Abbreviations: ADHD, attention-deficit/hyperactivity disorder; BH, Benjamini-Hochberg correction across the four outcomes; d, Cohen d for the difference in cyclic R² between participants with and without an informant-confirmed ADHD diagnosis; edf, effective degrees of freedom of the population cycle smooth; FDR, false discovery rate (participants whose within-person permutation P value survives FDR q < .05); GAMM, generalized additive mixed model. Robustly cyclical = cyclic R² or amplitude above the 95th percentile of a within-person permutation null (5,000 time shuffles). Cyclic R² = the cycle's share of a participant's within-person variance. Peak window = among robustly cyclical participants, the window (menstrual, luteal, or peri-ovulatory) with the participant's highest phase-elevation score, as in Figure 1B. Sample sizes differ by analysis: the population columns (participants, daily reports, edf, P, variance explained) use every rated day with cycle time from the 106 participants who had any (the analytic sample); the person-level columns (robustly cyclical, cyclic R², peak window, diagnosis contrast) use the 87 with adequate coverage of observed-onset cycle time on the ADHD scales. Sample sizes for every analysis are in eTable 1.
The two primary ADHD outcomes (inattention, hyperactivity-impulsivity) are listed first and the two secondary outcomes (negative affect, joint/muscle pain) below. Columns give the population GAMM (edf, P value, % of day-to-day variance); % robustly cyclical; cyclic R², the cycle's share of within-person variance (per-person, average and among those whose symptoms robustly cycled) as a measure of effect size; and the peak window (the percentage of robustly cyclical participants whose own curve peaked in each window; Figure 1B shows the same counts as shares of all 87 participants). Within-person associations of ovarian steroids with the four daily outcomes are reported in eTable 4.

### Person-level cyclicity

Individual trajectories varied widely (Figure 1A), and smoothing mixture models (75 participants; eMethods 4) recovered menstrual, luteal, and peri-ovulatory classes beside a flat class (inattention: flat 37, menstrual 22, luteal 16; hyperactivity-impulsivity: flat 40, peri-ovulatory 15, luteal 11, menstrual 9). Symptoms cycled robustly on at least 1 ADHD domain in 47 of 87 participants (54%; 95% CI, 44%-64%): 37 for inattention (43%; 33%-53%) and 23 for hyperactivity-impulsivity (26%; 18%-37%), 13 on both (Figure 2; Table 2; eTable 5).

**Figure 2.**
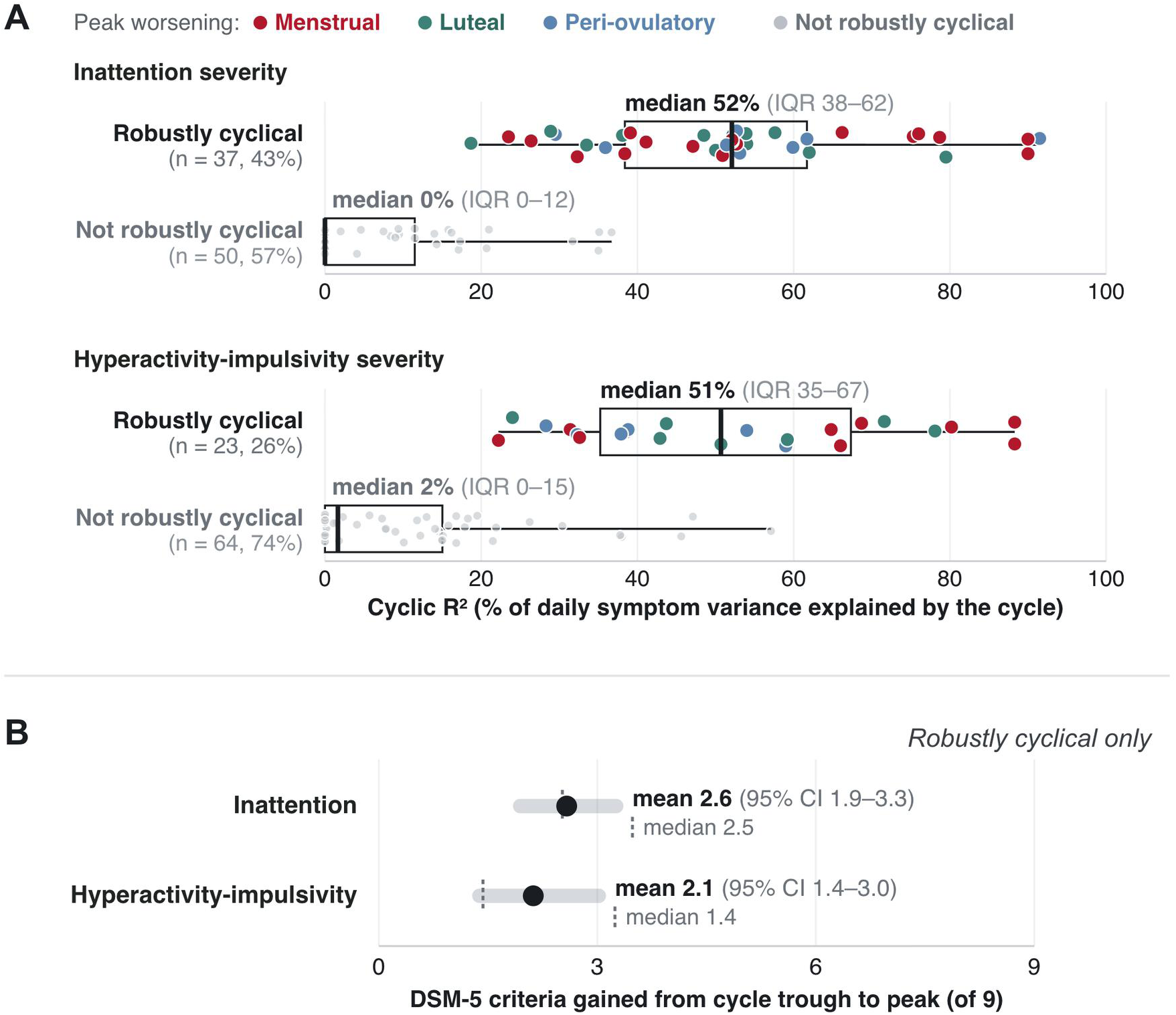
Size of the Cycle Effect in Participants With and Without Robust ADHD Cyclicity. (A) Per-person cyclic R² (the cycle’s share of each participant’s day-to-day symptom variance) for inattention and hyperactivity-impulsivity, in participants robustly cyclical versus not; each dot is one participant, colored by the window in which that participant’s own curve peaked (Figure 1B; gray = not robustly cyclical); boxes give the median and IQR and whiskers the range. (B) For robustly cyclical participants only, the mean model-implied swing (95% CI) in the number of DSM-5 criteria met between the cycle’s trough and peak (out of 9 per domain), with the median marked (dashed line). Robustly cyclical = cyclic R² or amplitude above the 95th percentile of a within-person permutation null (5,000 shuffles); because that null depends on each participant’s number of rated days, some participants who were not robustly cyclical have higher cyclic R² than some who were. Values for robustly cyclical participants are in-sample estimates in participants selected for exceeding their own null, so they are upper bounds; in 2-fold cross-validation, median cyclic R² among robustly cyclical participants whose two halves could both be evaluated was 17% (inattention) and 12% (hyperactivity-impulsivity; eMethods 3). N = 87 naturally cycling adults with ADHD symptoms. ADHD indicates attention-deficit/hyperactivity disorder; DSM-5, *Diagnostic and Statistical Manual of Mental Disorders*, Fifth Edition; IQR, interquartile range.

### Correlates of robust cyclicity

Robust ADHD cyclicity was more common in patients with informant-confirmed ADHD (exploratory; 69% vs 36%; odds ratio [OR], 3.9; 95% CI, 1.4-11.2; P = .004; q = .11 across 30 characteristics; eTable 6). Inattention cyclic R² was higher with informant-confirmed ADHD (d = 0.54; 95% CI, 0.10-0.98; P = .02; Table 2). No other demographic, comorbidity, or medication characteristic differed by either categorical or continuous cyclicity (Table 1; eTable 7). Restricting the sample to the 79 participants who met DSM-5 criteria by interview or informant report changed no prevalence by more than 1 percentage point (eTable 2).

### Menstrual-cycle risk windows

Among robustly cyclical participants, symptom elevation peaked most often in the menstrual window (inattention 16 of 37; hyperactivity-impulsivity 10 of 23), and otherwise in the luteal (13; 7) or peri-ovulatory (8; 6) windows (Figure 1B). The menstrual window ran from menses into the early follicular phase, not premenstrually.

### Where statistically robust, patient-level cycle effects were clinically relevant

In these participants, the cycle explained a median 52% (inattention) and 51% (hyperactivity-impulsivity) of daily symptom variance. In DSM-5 terms, the model-implied cycle swing in symptom criterion count (symptoms rated “often” or “very often” that day, 0–9 per domain) averaged 2.6 criteria for inattention (95% CI, 1.9-3.3) and 2.1 for hyperactivity-impulsivity (1.4-3.0; Figure 2; eMethods 3). Across both domains combined, the 47 participants cyclical on either domain swung by a mean of 3.5 criteria (95% CI, 2.6-4.5), and 12 (26%) by 5 or more.

### Working memory, but not inhibition, dipped where a patient’s own inattention cycle peaked (exploratory)

In 85 participants (321 in-lab visits), the working-memory deficit (higher = worse) was greater at the cycle positions where the participant’s own inattention cycle was high (within-person β = 0.27 per own-SD; 95% CI, 0.02-0.52; P = .03; q = .13; eFigure 2; eTable 8). Working memory did not track hyperactivity-impulsivity, and inhibitory-task performance tracked neither domain (eTable 8). Participants with greater menstrual worsening of inattention had poorer working memory just after menses (P = .06; Figure 1D) and, more strongly, at cycle positions where normative E2 is low (elevation × normative E2, β = −0.11; 95% CI, −0.17 to −0.04; P = .001), a pattern specific to E2 (joint P4 term, P = .95; eMethods 6).

### ADHD cyclicity frequently co-occurred with, yet was largely distinct from, cyclical affect and pain

At the patient level, many patients also showed robust cyclicity of negative affect (33%) and pain (33%). Cyclical worsening frequently co-occurred across domains (25 of 87 cycled in no domain, and 11 in all 3; Figure 3A). Next-day cross-lagged associations of ADHD and negative affect were similar in both directions (eTable 9). Most robustly cyclical participants remained so after within-person adjustment for same-day negative affect (inattention 22 of 37, median 79% of cyclic R² retained; hyperactivity-impulsivity 15 of 23, 76%; either domain, 32 of 47 [68%]; similar for previous-day, eTable 10; Figure 3B) or same-day pain (inattention 31 of 37; hyperactivity-impulsivity 16 of 23; either domain, 38 of 47 [81%]).

**Figure 3.**
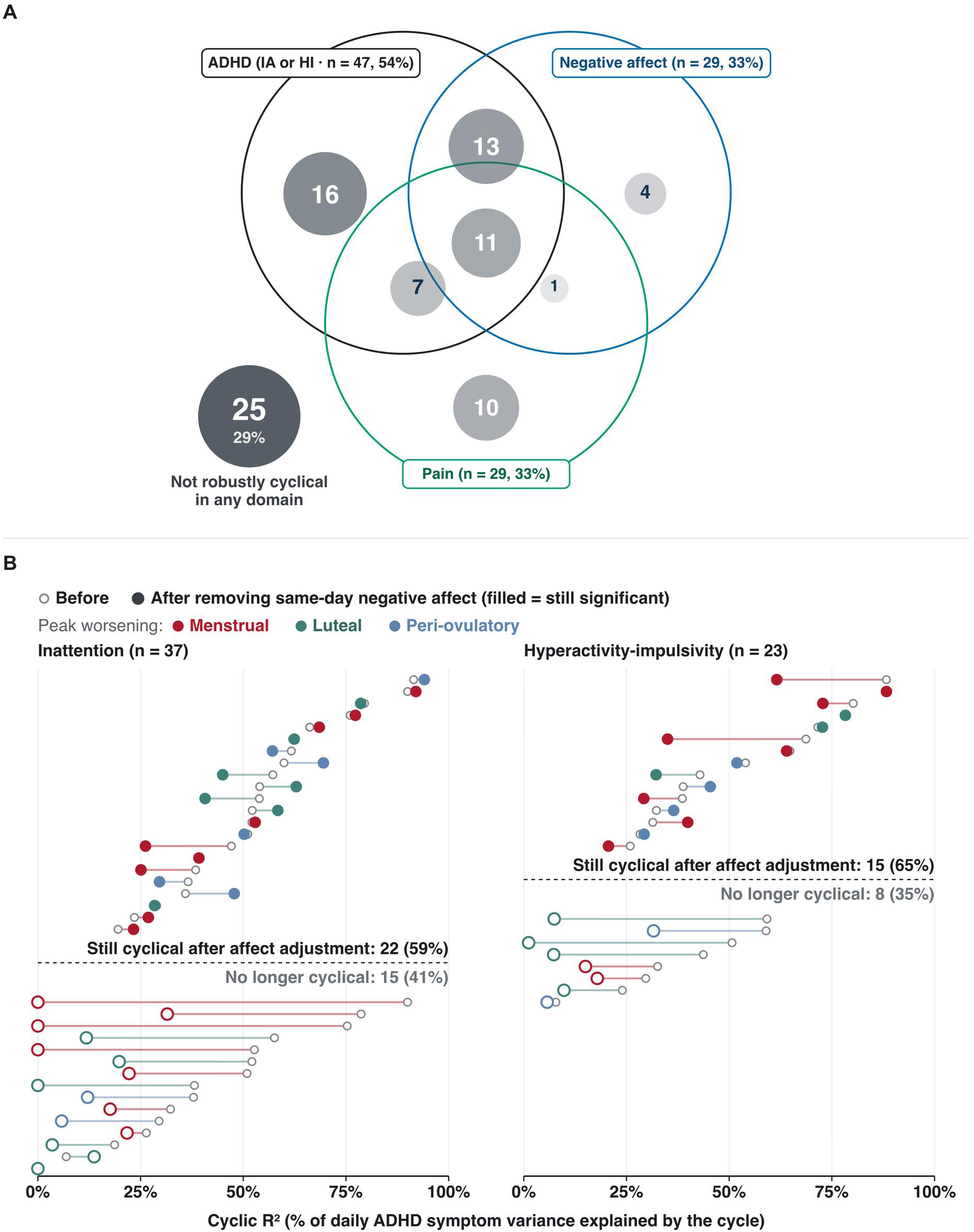
Co-occurrence of Robust Cyclicity Across Domains, and ADHD Cyclicity Before and After Adjustment for Same-Day Negative Affect. (A) Overlap of robust cyclicity among the 62 participants cyclical in ≥1 domain (ADHD = IA or HI); bubble size and shade are proportional to the number of participants in each region; 25 of 87 (29%) were cyclical in no domain. (B) For each participant with robust ADHD cyclicity, cyclic R² on the same days before (small open gray circle) and after (colored circle) same-day negative affect was removed within person; color = the window in which the participant’s own curve peaked (Figure 1B); filled = cyclicity still significant after adjustment (P < .05 against the participant’s own permutation null), shown above the dashed line; open = no longer significant. Inattention: 22 of 37 (59%) remained robustly cyclical; hyperactivity-impulsivity: 15 of 23 (65%). The median share of each participant’s cyclic R² retained was 79% (IQR, 20%-101%) and 76% (48%-100%), counting participants whose unadjusted refit did not reproduce their cyclicity as 0%; on the days with affect ratings, median cyclic R² fell from 52% to 28% and from 44% to 32% (52% and 51% on all days; eTable 10). N = 87 naturally cycling adults with ADHD symptoms. ADHD indicates attention-deficit/hyperactivity disorder; HI, hyperactivity-impulsivity; IA, inattention; IQR, interquartile range.

### Cyclical ADHD was not explained by PMDD

Although 39 of 78 (50%) endorsed retrospective PMDD criteria on the SCID-5 interview, no cycle met prospective C-PASS PMDD criteria (77 participants, 80 cycles; eTable 11), and retrospective PMDD report did not distinguish those with robust ADHD cyclicity (Table 1; eTable 12); retrospective and prospective PMDD status agreed poorly (κ = 0.05). Medication did not account for cyclicity: stimulant users had a flatter sample-average ADHD cycle (P ≤ .004; q = .02; eFigure 3) but were robustly cyclical on either ADHD domain at least as often as non-users (either domain, 63% vs 50%; P = .41), and the cycle did not differ by selective serotonin reuptake inhibitor use (eTable 13).

### Hormonal correlates of daily ADHD symptoms

Among robustly cyclical participants, lower within-person E2 and P4 accompanied worse hyperactivity-impulsivity (P = .004 and P < .001; q ≤ .03; inattention with E2, P = .07; eTable 4), and hormone slopes varied significantly across all participants (random-slope tests, P < .001 for inattention with E2 and P4 and hyperactivity-impulsivity with P4; eMethods 5). In the sample as a whole, neither E2, P4, nor their interaction predicted either ADHD symptom (eTable 4). E2 and P4 cycles did not differ detectably between those with and without robust ADHD cyclicity, in shape or level (all P ≥ .39; Figure 1C).

## Discussion

In this intensive longitudinal cohort, the sample-average cycle effect on ADHD symptoms was small and obscured clinically significant cyclicity in half of participants: in 46% of participants, symptoms were not robustly cyclical, while 54% showed robust cycle change in inattention or hyperactivity-impulsivity (a mean swing of 3.5 DSM-5 symptoms across both domains). Cyclicity was more common and larger with informant-confirmed ADHD (69% vs 36%), a finding not explained by greater symptom severity, which was similar with and without confirmation (eTable 2). A change of this size is highly significant for diagnostic assessment, because it could move a patient across the DSM-5 threshold of 5 symptoms (10). These effects emerged in naturally cycling individuals, were accompanied for inattention by parallel cyclical changes in objective working-memory performance, and were mostly not reducible to affective or pain changes.

Cyclical exacerbations fell into 3 patterns, each aligned with a DASH-MC hormone event (17). The most common ADHD pattern, worsening at and after menses when E2 is at its nadir, matches the framework’s hypothesized low or falling E2 risk window, grounded in experimental studies in which perimenstrual E2 administration (vs placebo) prevented menstrual declines in working memory (18) and steroid stabilization prevented mood worsening (29). Consistent with this window, working memory dipped at normatively low-E2 cycle positions in those with menstrual inattention worsening. A luteal worsening pattern was also common, potentially consistent with the luteal neurosteroid-surge sensitivity implicated in PMDD (30). A peri-ovulatory pattern, more common for hyperactivity-impulsivity, matched the E2-surge window the framework links to dysregulated reward-seeking (31) and substance use (32). Because the data are observational and mostly cover 1 cycle per person, these results establish within-person temporal association, not causal mechanism.

The most common worsening window was menstrual and postmenstrual. This is inconsistent with the concepts of PMDD and premenstrual exacerbation, because the postmenstrual window is when DSM-5 criteria require symptoms of PMDD, the only established cyclical psychiatric disorder, to be minimal or absent (10). A patient with robustly cyclical inattention can therefore screen negative on a premenstrual (luteal) mood assessment yet have clinically significant cycle-linked ADHD. Detecting this pattern required prospective daily ratings across the whole cycle, analyzed with flexible nonlinear curves and without assuming a window, rather than the premenstrual-versus-postmenstrual comparison required for DSM-5 PMDD or the research construct of premenstrual exacerbation (4). Notably, participants whose affect or inattention peaked at menses often endorsed PMDD retrospectively (8 of 10 and 11 of 15) but rarely showed the premenstrual elevation C-PASS requires (1 of 10 and 1 of 15; eTable 12), suggesting that they are aware of their cyclical pattern but may misplace its timing, perhaps mistaking menstrual for premenstrual worsening (33). Similarly, in a survey of 600 females with ADHD, most who reported cyclical change described luteal worsening (21), and surveys find provisional PMDD in 31% to 41% of females with ADHD (34); yet in our sample, half met retrospective SCID-5 PMDD criteria and no cycle met prospective criteria, suggesting that single-timepoint reports overstate PMDD in ADHD. Cyclical ADHD is therefore not PMDD misdiagnosed as ADHD; its worsening fell largely outside the premenstrual window. This supports the DASH-MC framework’s argument that cyclical vulnerability is a transdiagnostic (35) and multidimensional phenomenon, including separable luteal and menstrual sensitivities.

As most commonly observed in PMDD and other reproductive mood disorders (16, 36), the absence of hormone differences is consistent with differential sensitivity to normal hormone change rather than with endocrine or gynecologic abnormality, although salivary E2 immunoassays are limited by relatively high measurement error (37). Because measured hormones and cycle position move together, observational data cannot dissociate hormonal from other cycle-linked influences. Isolating hormonal mechanisms will require experimental manipulation, such as perimenstrual E2 stabilization (18), or ovarian suppression with a gonadotropin-releasing hormone agonist plus hormone add-back for luteal surge sensitivity (16), in patients with cyclical ADHD.

A central alternative explanation is that cyclical ADHD simply reflects undiagnosed comorbid PMDD. Two findings suggest that cyclical ADHD is not simply a consequence of cyclical mood. First, self-reported inattention cycles were accompanied by an objective working-memory signal at the same cycle positions, suggesting genuine cycle-linked cognitive change, and most patients remained robustly cyclical after within-person adjustment for same-day affect. Second, cross-lagged associations of similar size in both directions (eTable 9) argue against a one-way path from mood to attention; shared causes such as low menstrual E2 seem most plausible but require further confirmation (18).

The study has several clinical implications. First, in those with cyclical ADHD, a single diagnostic visit could fall on a cycle day when symptoms are unusually low or high, and it is unclear which should be regarded as ground truth (38). Second, screening only for cyclical mood change or PMDD would miss the 49% of those with cyclical ADHD symptoms whose affect did not cycle (Figure 3A); repeated assessment across the cycle appears to be needed (39). Third, whether cycle-timed treatment improves outcomes should be tested. Because stimulant effects appear sensitive to E2 and P4 (32), titrating dose to each patient’s worsening window, which was reported to be helpful and well tolerated in a small clinical series (20), is one candidate.

This study has limitations. The sample was predominantly White, young, and regularly cycling, limiting generalizability. Diagnostic criteria were based on a combination of methods (eMethods 1; eTable 2). Participants contributed a median of 1 cycle (mean 1.2), so person-level prevalence and peak-window estimates include substantial error, and cross-cycle stability remains to be tested. Medication was recorded once at baseline. The hyperactivity-impulsivity scale pools hyperactivity and impulsivity items, which may cycle differently. Missed diary days were probably lower-symptom days; assuming them 1 SD lower reduced cycle amplitudes by 11% to 44% but did not eliminate them (eTable 14). The median cyclic R² and DSM-5 swing are upper bounds, because each was estimated on the same days used to classify participants as robustly cyclical; when each half of a participant’s days was predicted from the other half, the median cyclic R² was 17% (inattention) and 12% (hyperactivity-impulsivity; eMethods 3).

This study also has strengths. Cycle position was anchored to observed menses onsets and to ovulation coded from daily hormone patterns, then standardized with PACTS; cyclicity was tested within each person against that person’s own permutation null rather than inferred from group averages; PMDD was assessed prospectively with the standard C-PASS rather than by recall; and objective cognition was measured. The sample is also larger than that underlying the only community-based estimate of premenstrual exacerbation of depression, which rested on 58 participants with depressive disorders (11).

Priorities for future work include prospective multi-cycle studies of within-person stability and trials of cycle-timed treatment; if replicated, menstrual-cycle sensitivity should be considered as a potential specifier or modifier of symptoms in ADHD.

## Data Availability

Deidentified daily-diary, hormone, and lab-visit data have been deposited in the NIMH Data Archive (collection for R01MH119119) and will be available to qualified investigators under NDA data-use terms on publication. Analysis code is available from the corresponding author on reasonable request.

## Article Information

### Author Contributions

Dr Eisenlohr-Moul had full access to all of the data in the study and takes responsibility for the integrity of the data and the accuracy of the data analysis. Concept and design: Eisenlohr-Moul, Peters, Schmalenberger, Martel. Acquisition, analysis, or interpretation of data: All authors. Drafting of the manuscript: Eisenlohr-Moul. Critical review of the manuscript for important intellectual content: All authors. Statistical analysis: Eisenlohr-Moul, Ross, Nagpal. Obtained funding: Martel, Eisenlohr-Moul. Administrative, technical, or material support: Knapp, Nagpal, Patterson, Schmalenberger. Supervision: Eisenlohr-Moul, Martel.

### Additional Information

Claude Code (Anthropic; Claude 4 and 5 family models; May– November 2026) was used under the authors’ direction to generate R code that converts our raw model outputs into tables and figures, creating replicable code for these outputs. The same AI tools identified redundant or low-priority text for removal or relocation to the supplement and reviewed and updated labels for figures, tables, and e-content in the manuscript to ensure proper ordering and accuracy across the text. A double-human-review process checked all AI-formatted content against raw outputs; no errors were found. In sum, the authors verified all content and take full responsibility for it.

## REFERENCES

1. Faraone SV, Banaschewski T, Coghill D, et al.: The World Federation of ADHD International Consensus Statement: 208 evidence-based conclusions about the disorder. Neurosci Biobehav Rev 2021; 128:789–818

2. Eng AG, Nirjar U, Elkins AR, et al.: Attention-deficit/hyperactivity disorder and the menstrual cycle: theory and evidence. Horm Behav 2024; 158:105466

3. Kooij JJS, de Jong M, Agnew-Blais J, et al.: Research advances and future directions in female ADHD: the lifelong interplay of hormonal fluctuations with mood, cognition, and disease. Front Glob Womens Health 2025; 6:1613628

4. Eisenlohr-Moul TA, Girdler SS, Schmalenberger KM, et al.: Toward the reliable diagnosis of DSM-5 premenstrual dysphoric disorder: the Carolina Premenstrual Assessment Scoring System (C-PASS). Am J Psychiatry 2017; 174:51–59

5. Reilly TJ, Patel S, Unachukwu IC, et al.: The prevalence of premenstrual dysphoric disorder: systematic review and meta-analysis. J Affect Disord 2024; 349:534–540

6. Armour M, Parry K, Manohar N, et al.: The Prevalence and Academic Impact of Dysmenorrhea in 21,573 Young Women: A Systematic Review and Meta-Analysis. J Womens Health 2019; 28:1161–1171

7. Wang L, Yan Y, Qiu H, et al.: Prevalence and Risk Factors of Primary Dysmenorrhea in Students: A Meta-Analysis. Value Health 2022; 25:1678–1684

8. Kuehner C, Nayman S: Premenstrual exacerbations of mood disorders: findings and knowledge gaps. Curr Psychiatry Rep 2021; 23:78

9. Gehlert S, Song IH, Chang C-H, et al.: The prevalence of premenstrual dysphoric disorder in a randomly selected group of urban and rural women. Psychol Med 2009; 39:129–136

10. American Psychiatric Association: Diagnostic and Statistical Manual of Mental Disorders. DSM-5-TR. American Psychiatric Association Publishing, 2022

11. Hartlage SA, Brandenburg DL, Kravitz HM: Premenstrual exacerbation of depressive disorders in a community-based sample in the United States. Psychosom Med 2004; 66:698–706

12. Solberg BS, Halmøy A, Engeland A, et al.: Gender differences in psychiatric comorbidity: a population-based study of 40 000 adults with attention deficit hyperactivity disorder. Acta Psychiatr Scand 2018; 137:176–186

13. Schiller CE, Johnson SL, Abate AC, et al.: Reproductive steroid regulation of mood and behavior. Compr Physiol 2016; 6:1135–1160

14. Hampson E: Estrogens, aging, and working memory. Curr Psychiatry Rep 2018; 20:109

15. Schmidt PJ, Martinez PE, Nieman LK, et al.: Premenstrual dysphoric disorder symptoms following ovarian suppression: triggered by change in ovarian steroid levels but not continuous stable levels. Am J Psychiatry 2017; 174:980–989

16. Wei SM, Wakim P, Martinez PE, et al.: Differential effects of ovarian steroids in women with and without premenstrual dysphoric disorder. Am J Psychiatry 2025; 182:922–934

17. Peters JR, Schmalenberger KM, Eng AG, et al.: Dimensional Affective Sensitivity to Hormones across the Menstrual Cycle (DASH-MC): a transdiagnostic framework for ovarian steroid influences on psychopathology. Mol Psychiatry 2025; 30:251–262

18. Schmalenberger KM, Mulligan EM, Barone JC, et al.: Effects of acute estradiol administration on perimenstrual worsening of working memory, verbal fluency, and inhibition in patients with suicidal ideation: a randomized, crossover clinical trial. Psychiatry Res 2024; 342:116188

19. Roberts B, Eisenlohr-Moul T, Martel MM: Reproductive steroids and ADHD symptoms across the menstrual cycle. Psychoneuroendocrinology 2018; 88:105–114

20. de Jong M, Wynchank DSMR, van Andel E, et al.: Female-specific pharmacotherapy in ADHD: premenstrual adjustment of psychostimulant dosage. Front Psychiatry 2023; 14:1306194

21. Osianlis E, Thomas EHX, Li Q, et al.: ADHD in females: survey findings on symptoms across hormonal life stages. J Psychiatr Res 2026; 193:208–215

22. Zaritsky R, Reed SC, Evans SM: Changes in ADHD symptoms and mood across the menstrual cycle in females treated with stimulants: a pilot study. J Atten Disord 2026; 30:329–341

23. Barkley RA: Barkley Adult ADHD Rating Scale-IV (BAARS-IV). Guilford Press, 2011

24. Schmalenberger KM, Tauseef HA, Barone JC, et al.: How to study the menstrual cycle: practical tools and recommendations. Psychoneuroendocrinology 2021; 123:104895

25. Nagpal A, Schmalenberger KM, Barone JC, et al.: How to study the menstrual cycle as a continuous variable: implementing phase-aligned cycle time scaling (PACTS) with the menstrualcycleR package. Psychoneuroendocrinology 2025; 181:107584

26. Endicott J, Nee J, Harrison W: Daily Record of Severity of Problems (DRSP): reliability and validity. Arch Womens Ment Health 2006; 9:41–49

27. Benjamini Y, Hochberg Y: Controlling the false discovery rate: a practical and powerful approach to multiple testing. J R Stat Soc Series B 1995; 57:289–300

28. Phipson B, Smyth GK: Permutation P-values should never be zero: calculating exact P-values when permutations are randomly drawn. Stat Appl Genet Mol Biol 2010; 9:Article39

29. Eisenlohr-Moul TA, Bowers SM, Prinstein MJ, et al.: Effects of acute estradiol and progesterone on perimenstrual exacerbation of suicidal ideation and related symptoms: a crossover randomized controlled trial. Transl Psychiatry 2022; 12:528

30. Martinez PE, Rubinow DR, Nieman LK, et al.: 5α-Reductase inhibition prevents the luteal phase increase in plasma allopregnanolone levels and mitigates symptoms in women with premenstrual dysphoric disorder. Neuropsychopharmacology 2016; 41:1093–1102

31. Jacobs E, D’Esposito M: Estrogen shapes dopamine-dependent cognitive processes: implications for women’s health. J Neurosci 2011; 31:5286–5293

32. Justice AJ, de Wit H: Acute effects of d-amphetamine during the follicular and luteal phases of the menstrual cycle in women. Psychopharmacology (Berl) 1999; 145:67–75

33. McFarland C, Ross M, DeCourville N: Women’s theories of menstruation and biases in recall of menstrual symptoms. J Pers Soc Psychol 1989; 57:522–531

34. Broughton T, Lambert E, Wertz J, et al.: Increased risk of provisional premenstrual dysphoric disorder (PMDD) among females with attention-deficit hyperactivity disorder (ADHD): cross-sectional survey study. Br J Psychiatry 2025; 226:410–417

35. Zhou J, Muse Z, Bränn E, et al.: Bidirectional association between premenstrual disorders and psychiatric disorders. JAMA Netw Open 2026; 9:e2611765

36. Schmidt PJ, Nieman LK, Danaceau MA, et al.: Differential behavioral effects of gonadal steroids in women with and in those without premenstrual syndrome. N Engl J Med 1998; 338:209–216

37. Arslan RC, Blake K, Botzet LJ, et al.: Not within spitting distance: salivary immunoassays of estradiol have subpar validity for predicting cycle phase. Psychoneuroendocrinology 2023; 149:105994

38. Sibley MH, Arnold LE, Swanson JM, et al.: Variable patterns of remission from ADHD in the Multimodal Treatment Study of ADHD. Am J Psychiatry 2022; 179:142–151

39. Eisenlohr-Moul TA: Toward precision reproductive psychiatry: a call for targeted, transdiagnostic approaches to premenstrual disorders. Am J Psychiatry 2025; 182:894–897

40. Wactawski-Wende J, Schisterman EF, Hovey KM, et al.: BioCycle study: design of the longitudinal study of the oxidative stress and hormone variation during the menstrual cycle. Paediatr Perinat Epidemiol 2009; 23:171–184

