## Supplementary Material for "Menstrual Cycle Exacerbation of ADHD Symptoms in Adults: Common, Person-Specific, and Not Confined to the Luteal Phase"

Contents (the STROBE checklist is a separate file)

eMethods 1. Samples and inclusion
eMethods 2. Population cycle models
eMethods 3. Person-level cyclicity
eMethods 4. Peak windows
eMethods 5. Hormones
eMethods 6. Objective cognition
eMethods 7. Prospective and retrospective PMDD
eMethods 8. Comorbidity, medication, and continuous analyses
eMethods 9. Missing data
eTable 1. Sample definitions and sample size for each analysis
eTable 2. Person-level results restricted to participants meeting DSM-5 criteria by interview or informant report
eTable 3. Within-person reliability of the daily composites
eTable 4. Daily hormones and symptoms
eTable 5. Co-occurrence of robust cyclicity across domains
eTable 6. Robust cyclicity by informant-confirmed ADHD
eTable 7. Participant characteristics and cyclicity as a continuous exposure
eTable 8. Cognitive performance aligned with each participant's symptom cycle
eTable 9. Next-day cross-lagged associations of ADHD symptoms and negative affect
eTable 10. ADHD cyclicity after adjustment for negative affect or pain
eTable 11. Prospective PMDD (C-PASS)
eTable 12. Retrospective PMDD report: items, and agreement with daily ratings by peak window
eTable 13. Medication and cyclicity
eTable 14. Diary nonresponse and missing-data sensitivity
eFigure 1. Participant flow
eFigure 2. Working memory aligned with each participant's inattention cycle
eFigure 3. Sample-average cycle by stimulant and SSRI use

### eMethods

### eMethods 1: Samples and inclusion

Of 125 participants with daily data, 10 were excluded before analysis: 9 who met neither DSM-5 route (interview or informant) and either rated daily ADHD symptoms at zero on most days, reported no ADHD symptoms on the SCID-5, or had no diagnostic data and daily ratings in the lowest quarter on both scales; and 1 with too few cycle anchors. The analytic sample (n = 106) is everyone with at least 1 rated day with cycle time, and population models used all such days. Person-level analyses used only days anchored by an observed menses onset and required at least 10 such days with at least 1 in each quarter of the cycle (3 quarters for 1 participant); 87 participants met this rule for the daily ADHD ratings. Four others met it only for a daily cognitive task not analyzed here and contribute to population models only. eTable 1 lists each sample.

Diagnosis contrasts use the 81 of the 87 with an informant-adjudicated diagnosis (42 confirmed, 39 not), made by a licensed clinical psychologist. Four participants rated pain at the scale floor every day; with nothing to test, their pain was classified as non-cyclical (pain cyclicity, 29 of 87).

Complete cycles ranged from 20 to 37 days. Five cycles in 5 participants fell outside 21 to 35 days; each had a coded ovulation and was kept, and excluding these participants changed no prevalence by more than 2 percentage points. A sixth out-of-range cycle with no coded ovulation was dropped. Cycle length did not differ by cyclicity (all $P > .06$).

The 87 did not differ from the 38 other participants with daily data in age, race, or stimulant use ($P \geq .12$) and had slightly more SCID-5 symptoms (12.8 vs 10.7; $P = .05$).

**Cycle anchors and imputation.** Cycle time was computed with pacts_scaling in menstrualcycleR 1.1.0 (eReference 1). A missing ovulation in a 21- to 35-day cycle was imputed 15 days before the next onset; a confirmed ovulation without a closing onset received an onset 14 days later; and days before the first onset were placed by imputing ovulation 15 days before it. For population models only, days after the last onset were placed using the participant's median cycle length (otherwise the cycle length reported at screening, otherwise 28 days).

### eMethods 2: Population cycle models

Population cycle effects (Table 2) were estimated with generalized additive mixed models (eReferences 2 and 3): a population cyclic smooth of PACTS time, with a random intercept and a random cyclic smooth for each participant, fit to every rated day with cycle time (106 participants; about 2,580 days per outcome). We compared 6 random-effects structures. Structures with cycle-level terms fit better by AIC, but only 24 participants had enough coverage in 2 cycles to estimate them, so we used the per-person cyclic smooth for all outcomes; the cycle effect was $P < .001$ for every outcome under every structure. Families were chosen from residual diagnostics: Tweedie (log link) for the ADHD severities and gamma (log link) for negative affect and pain. Trough-to-peak swings were 12% (inattention), 16% (hyperactivity-impulsivity), 16% (negative affect), and 18% (pain) of the outcome mean. Benjamini-Hochberg correction was applied within each family of tests, and we report uncorrected and corrected P values.

### eMethods 3: Person-level cyclicity

**The curve.** For each participant and outcome, we fit a cyclic cubic spline of PACTS time (idiographic_gam 1.0.7; basis dimension 10, penalized) to all of that participant's days, on the outcome's own scale. Because most participants contributed about 1 cycle, we fit one pooled curve per person, adding a cycle-specific intercept only where 2 cycles each covered at least 3 quarters (14 participants per ADHD scale). Only 2 participants had all 4 quarters in 2 cycles, so cross-cycle stability could not be tested.

**Effect sizes.** Cyclic $R^2$ is the share of the participant's day-to-day variance explained by the curve. It is nearly unrelated to the participant's mean level ($|r| \leq 0.05$ for the ADHD and affect outcomes; 0.25 for pain). Amplitude is the curve's peak minus trough in outcome units. The two rank participants almost identically (Spearman $\rho$, 0.91 to 0.96).

**Detection.** A participant was robustly cyclical on an outcome if cyclic $R^2$ or amplitude exceeded the 95th percentile of the same statistic across 5,000 within-person time shuffles of their own days ($P = [b + 1]/[m + 1]$). Cyclic $R^2$ and amplitude alone selected nearly the same participants ($\kappa$, 0.90 to 0.97); combining them added at most 1 participant per outcome to the cyclic $R^2$ rule. Table 1 contrasts were repeated with cyclic $R^2$ and amplitude as continuous exposures (eTable 7).

**Calibration and power.** Shuffling days also removes serial dependence, so the null could be too permissive. We therefore injected cyclic signals of known size into the day schedules of 78 real participants. With no signal, the cyclic $R^2$ criterion flagged 4.7% of 468 tests (the combined rule can only flag more and was not simulated). Power was 21%, 34%, 59%, 80%, and 89% at a true cyclic $R^2$ of 0.1, 0.2, 0.3, 0.4, and 0.5; at 0.3, it was 28% with fewer than 20 rated days, 65% with 20 to 29, and 91% with 30 or more. Benjamini-Hochberg correction across participants within each outcome left 19 (inattention), 4 (hyperactivity-impulsivity), 7 (negative affect), and 19 (pain) participants significant at $q < .05$ (Table 2).

In 2-fold cross-validation (curve fit to half of a participant's days and evaluated on the other half), cyclic $R^2$ stayed positive for 16 of 25 robustly cyclical participants on inattention and 13 of 19 on hyperactivity-impulsivity, but for fewer on negative affect (11 of 25) and pain (4 of 17). In these participants, median cross-validated cyclic $R^2$ was 17% (inattention) and 12% (hyperactivity-impulsivity), versus 55% and 52% in-sample, so the in-sample medians in the main text are upper bounds.

**DSM-5 count swing.** The daily DSM-5 count (symptoms rated often or very often, 0 to 9 per domain) was modeled with the same per-person model; the swing is the model-implied peak minus trough. Among robustly cyclical participants, the mean swing was 2.6 inattention symptoms (95% CI, 1.9 to 3.3; median 2.5; n = 37) and 2.1 hyperactivity-impulsivity symptoms (1.4 to 3.0; median 1.4; n = 23); CIs are percentile bootstrap intervals (10,000 resamples). The combined swing adds the two domain curves before taking peak minus trough; among the 47 cyclical on either domain, it averaged 3.5 (2.6 to 4.5; median 2.9), and 12 (26%) swung by 5 or more.

### eMethods 4: Peak windows

**Mixture models.** Smoothing mixture models (Nagpal and colleagues, in press) were fit to the 75 participants with at least 5 rated days in each half of the cycle (log[y + 1] centered on each participant's mean; per-person random intercept and slope within class; 2 to 6 classes). Information criteria kept favoring more classes, so we chose the number by inspecting the solutions and named each class by where its mean curve peaked. Inattention: flat 37, menstrual 22, luteal 16. Hyperactivity-impulsivity: flat 40, peri-ovulatory 15, luteal 11, menstrual 9. Negative affect: flat 38, luteal 22, peri-ovulatory 15. Pain: flat 47, luteal 17, menstrual 11. The classes are descriptive.

**Phase-elevation scores.** Each participant was scored on 3 a priori windows on the PACTS axis (menses = 0; ovulation = ±1): luteal ($-0.75 < t \le -0.1$), menstrual ($-0.1 < t \le 0.45$, menses through the early follicular phase), and peri-ovulatory ($t \ge 0.75$). The score is the mean deviation of the participant's curve from their cycle mean within the window, divided by their daily SD. A robustly cyclical participant's peak window is the window with the highest score (Table 2; Figure 1B). For the Figure 1D working-memory model only, the menstrual window was the interquartile range of peak times in the menstrual inattention class (+0.12 to +0.39).

### eMethods 5: Hormones

**Assays.** Morning saliva was assayed in duplicate for E2 (Salimetrics 1-3702; range, 0.5 to 32 pg/mL) and P4 (Salimetrics 1-1502; range, 3.33 to 2,430 pg/mL; diluted 2-fold) by enzyme immunoassay, and for luteinizing hormone by electrochemiluminescence (Meso Scale Discovery K1516FK; range, 2.74 to 6,825 pg/mL), on 74 plates. Manufacturer intra- and inter-assay coefficients of variation were 6.3% to 8.1% and 6.0% to 8.9% for E2 and 4% to 8% and 6% to 10% for P4. Values below range were kept as estimates, and wells with high replicate error were rerun. Salivary E2 immunoassays have limited validity (eReference 4), which would attenuate E2 associations.

**Daily hormone models (eTable 4).** (A) Each outcome was regressed on the within-person 5-day rolling hormone level, with the person mean as a covariate and a random intercept. In robustly cyclical participants, lower E2 and P4 accompanied worse hyperactivity-impulsivity (n = 23; $P = .004$ and $P < .001$), and lower E2 accompanied worse inattention at $P = .07$ (n = 37). Likelihood-ratio tests of random slopes showed that hormone slopes varied across participants for inattention with E2 ($\chi^2 = 32.7$) and P4 ($\chi^2 = 35.1$) and for hyperactivity-impulsivity with P4 ($\chi^2 = 33.8$; all $P < .001$), but not for hyperactivity-impulsivity with E2 ($P = .28$); average slopes were near zero ($P \ge .24$). (B) In mixed models of person-standardized E2, P4, and their product, with each outcome's family and random E2 and P4 slopes, no term predicted any outcome ($P \ge .05$). (C) A tensor-product E2 × P4 surface predicted negative affect and pain ($P \le .006$) but neither ADHD domain in the form matched to panel B ($q \ge .19$). (D) Previous-day decline in rolling P4 was associated with worse negative affect ($P = .01$); E2 change was not associated with any outcome. These models were not adjusted for cycle time, which indexes the same hormone sequence; with it included, the P4 associations were no longer significant.

**Hormone cycles by cyclicity (Figure 1C).** Cyclic mixed models of raw E2 and P4 with a difference smooth by cyclicity status (adjusted for age and BMI; 74 participants) found no difference ($P = .63$ for E2 and .86 for P4). The data are compatible with differences of up to ±13% of mean E2 and ±42% of mean P4 (90% pointwise band). Using each participant's larger ADHD cyclic $R^2$ as a continuous moderator gave the same result ($P = .59$ and .15).

### eMethods 6: Objective cognition

**Battery.** Participants completed an in-lab battery at up to 4 cycle-scheduled visits; 85 of the 87 had visits at a known cycle position (311 to 321 visits per model). Tasks were z-scored and averaged into composites scored so that higher = worse, winsorized at 2.5 SD: working memory (n-back d′, symmetry span, letter-number sequencing; eReferences 5-7) and inhibition (antisaccade reaction time, Stroop interference; eReferences 8 and 9). The stop-signal task was dropped because it correlated negatively with the other inhibition tasks. Visits scheduled for a given phase spanned nearly the whole cycle on the PACTS axis, so every analysis used each visit's actual PACTS position. The working-memory tasks correlated between people (mean $r = 0.36$) but only weakly within person (mean $r = 0.10$), so each task is also reported separately.

**Alignment with each participant's inattention cycle (eTable 8; eFigure 2).** Each participant's inattention cycle was summarized as 1 harmonic (cosine and sine) from a mixed model of the daily ratings with random harmonic coefficients, scaled by the participant's daily SD. Predicted inattention at each lab visit was split into the participant's mean across visits and the visit's deviation from that mean. The composite was regressed on both parts, adjusting for the population cycle, practice (a smooth of session number), and a person random intercept. Working memory was worse where the participant's own inattention cycle was high (within-person $\beta = 0.27$ per own-SD; SE, 0.13; $P = .03$; $q = .13$ across 4 pairings), in the same direction for each task ($\beta$, 0.15 to 0.20; $P = .21$ to .36). The other pairings were null ($P \geq .39$). With unshrunk per-person coefficients, the slope was 0.10 ($P = .13$). This analysis is exploratory.

**Working memory and normative E2 (Figure 1D).** Each visit was assigned the normative serum E2 value at its cycle position from a cyclic model of BioCycle Study data (eReference 10) on the same PACTS axis. The composite was regressed on the participant's menstrual inattention elevation score, normative E2, and their product, with practice and a person random intercept. Participants with greater menstrual elevation had poorer working memory where normative E2 was low ($\beta = -0.11$ per SD of each; 95% CI, −0.17 to −0.04; $P = .001$; on the a priori window, $P < .001$). The effect was specific to E2: with both products in one model, the E2 product was unchanged ($\beta = -0.11$; $P = .009$) and the P4 product was null ($\beta = 0.00$; $P = .95$). Because this model fixes the cycle shape to the normative E2 curve, we also fit a model with a free cyclic shape, which gave a weaker trend ($P = .06$); across its 6 a priori window-by-composite cells, no window-matched effect was found. This analysis is exploratory.

### eMethods 7: Prospective and retrospective PMDD

**Scoring.** The Carolina Premenstrual Assessment Scoring System (eReference 11) was applied to the daily DRSP: premenstrual days −7 to −1 vs postmenstrual days 4 to 10, at least 3 ratings per window, and an elevation of at least 30% of the item's scale range. Each participant was scored with menses or ovulation anchoring, whichever gave more scorable cycles (77 participants; 80 cycles). No cycle met PMDD criteria; 8 cycles met menstrually related mood disorder criteria and 3 showed premenstrual exacerbation (eTable 11). Only 3 participants had 2 scorable cycles, so results are reported per cycle. The premenstrual elevations that did occur were mainly physical (physical symptoms, fatigue, sleep, appetite).

**Retrospective report.** Thirty-nine of 78 participants (50%) met provisional PMDD criteria on the SCID-5, which did not distinguish participants with robustly cyclical ADHD (Table 1). No SCID-5 PMDD item was associated with ADHD cyclicity after correction (eTable 12); endorsing premenstrual difficulty concentrating did not identify cyclical inattention (37% of endorsers vs 36% of non-endorsers).

**Retrospective vs prospective report by peak window (eTable 12).** Among participants robustly cyclical on negative affect, those with a menstrual peak endorsed retrospective PMDD in 8 of 10 but met the C-PASS premenstrual elevation in only 1 of 10; luteal peakers did so in 56% and 63% (C-PASS, menstrual vs luteal, P = .04). For inattention, the figures were 73% and 7% (menstrual) and 38% and 63% (luteal; P = .009). Retrospective and prospective status agreed poorly overall (κ = 0.05; 69 participants).

### eMethods 8: Comorbidity, medication, and continuous analyses

**Comorbidity.** Diagnoses were coded from the SCID-5 Clinical Version. SCID-5 module data were missing for 9 of the 87, so comorbidity and retrospective PMDD comparisons use 78. No diagnosis differed by cyclicity (Table 1). Of the 30 characteristics in Table 1, only informant-confirmed ADHD differed at P < .05 (67% vs 34%; OR, 3.9; P = .004; q = .11).

**Medication (eTable 13; eFigure 3).** Medication was recorded once, at baseline, so it could not enter the within-person tests. We refit each population model with a medication-by-cycle difference smooth. The cycle did not differ by SSRI use (P = .26 to .84). Stimulant users had a flatter sample-average cycle for inattention (P = .004), hyperactivity-impulsivity (P = .003), and negative affect (P = .02), but not pain. Stimulant users were nonetheless robustly cyclical at least as often as non-users (either ADHD domain, 63% vs 50%; P = .41), which suggests that the flatter average reflects individual cycles peaking at different times. Excluding stimulant users, 36% (inattention), 28% (hyperactivity-impulsivity), and 50% (either domain) were robustly cyclical; excluding SSRI users, 35%, 22%, and 46%. With 15 to 23 users and no dosing records, these analyses show only that medication did not produce the cyclicity.

**Informant-confirmed diagnosis (eTable 6).** Robust cyclicity was compared by informant-confirmed diagnosis with Fisher exact tests (81 participants; BH across 5 domains). Unconfirmed participants who met the SCID-5 symptom criterion had interview and daily symptom levels similar to those of confirmed participants (eTable 2).

**Continuous analyses (eTable 7).** Table 1 contrasts were repeated with the larger of each participant's 2 ADHD cyclic $R^2$ values (and, separately, amplitude) as a standardized continuous exposure, using logistic models, Spearman correlations, or linear slopes, with BH correction within family. Across 46 contrasts, the DSM-5 count swing rose with cyclic $R^2$ (P < .001), informant-confirmed ADHD had an OR of 1.9 per SD (P = .01; q = .11), and hormone-sampling days decreased with cyclicity (ρ = −0.34; q = .006). No other contrast reached q < .10.

### eMethods 9: Missing data

Diary nonresponse varied across the cycle (P < .001), peaking around menses. In a mixed logistic model, participants were less likely to skip the day after a higher-symptom day, significantly only for inattention (P = .02), so missed days were, if anything, lower-symptom days (eTable 14A). In a pattern-mixture analysis that imputed missed days and shifted them 1 SD lower, the population cycle amplitude fell by 11% (hyperactivity-impulsivity) to 44% (pain), but no cycle effect disappeared (eTable 14B). Per-person cyclicity of missingness was not correlated with symptom cyclicity (ρ = 0.11; P = .34).

### eTables

### eTable 1: Sample definitions and sample size for each analysis

#### (A) Sample definitions.

| N | Definition |
|---|---|
| 125 | Provided any daily diary data |
| 115 | Remained after the 10 exclusions made before any analysis (eMethods 1) |
| 91 | Adequate cycle coverage on ≥1 daily measure (the 4 symptom ratings or the 2 daily cognitive games) |
| 87 | Person-level sample: adequate coverage of observed-onset cycle time on inattention or hyperactivity-impulsivity (per-person testing) |
| 81 | Person-level sample with an informant-adjudicated diagnosis |
| 106 | Analytic sample (population models): every rated day with cycle time, including days after the last observed onset (Table 2) |

Of 125 participants with daily data, 10 were excluded before analysis (eMethods 1). Of the remaining 115, 11 had no day on observed anchors (2 of these had imputed cycle time and enter population models only, giving the analytic sample of 106), 11 had fewer than 10 scalable days, and 2 lacked a day in at least 1 cycle quarter (a third with 3 quarters was admitted), leaving 91. Four had adequate coverage only on a daily cognitive task not analyzed here, leaving 87 for person-level analyses, of whom 81 had an informant-adjudicated diagnosis.

#### (B) Sample size for each analysis.

| Analysis | Participants (days or visits) | Where reported |
|---|---|---|
| Population-average cycle models | 106 (2,571-2,583 rated days per outcome) | Table 2 |
| Per-person cyclicity (detection, cyclic $R^2$, amplitude, peak window) | 87 (pain: 87, including 4 who never reported pain, counted not cyclical) | Table 2; Figures 1A, 1B, and 2; eTable 5 |
| Per-person affect and pain adjustment | Robustly cyclical only: 37 (inattention) and 23 (hyperactivity-impulsivity); previous-day affect 35 and 22 | Figure 3B; eTable 10 |
| Next-day cross-lagged associations | 109 (2,235 consecutive-day pairs; every rated day, with or without a cycle position) | eTable 9 |
| Within-person hormone models (levels, day-to-day change, E2 × P4) | 96-97 (2,152-2,360 days) | eTable 4 |
| Hormone cycles by robust ADHD cyclicity | 74 (79 of the 87 had hormone data; 5 lacked age or BMI) | Figure 1C; eMethods 5 |
| In-lab cognition | 85 (321 working-memory and 314 inhibition visits) | Figure 1D; eTable 8; eMethods 6 |
| Smoothing mixture models | 75 | eMethods 4 |
| Informant-confirmed diagnosis contrasts | 81 (42 confirmed, 39 not) | Table 1; eTable 6 |
| Diagnostic restriction | 79 meeting DSM-5 criteria by interview or informant | eTable 2 |
| Medication and the population cycle | 92 (stimulants: 23 users, 69 non-users); 93 (SSRIs: 19 users, 74 non-users) | eTable 13; eFigure 3 |
| Medication and per-person cyclicity | 80 with a stimulant record (16 users); 78 with an SSRI record (15 users) | Table 1; eTable 13 |
| Prospective PMDD (C-PASS) | 77 of the 87 with ≥1 scorable cycle (80 cycles), each scored under the anchoring that gave that participant more scorable cycles | Table 1; eTable 11; eMethods 7 |
| Diary nonresponse | 107 (2,549-2,561 person-days) | eMethods 9; eTable 14 |
| Retrospective SCID-5 PMDD report | 78 with SCID-5 module data (69 of them also with a scorable C-PASS cycle) | Table 1; eTable 12 |

Each analysis used every participant who met its data requirement.

### eTable 2: Person-level results restricted to participants meeting DSM-5 criteria by interview or informant report

| Quantity | Full sample (87 with per-person tests) | Restricted to those meeting DSM-5 criteria by interview or informant (79 with per-person tests) |
|---|---|---|
| Robustly cyclical: Inattention | 37 of 87 (43%) | 33 of 79 (42%) |
| Median cyclic $R^2$ among cyclical: Inattention | 52% | 51% |
| Robustly cyclical: Hyperactivity-impulsivity | 23 of 87 (26%) | 20 of 79 (25%) |
| Median cyclic $R^2$ among cyclical: Hyperactivity-impulsivity | 51% | 43% |
| Robustly cyclical: Negative affect | 29 of 87 (33%) | 27 of 79 (34%) |
| Robustly cyclical: Joint/muscle pain | 29 of 87 (33%) | 26 of 79 (33%) |
| Robustly cyclical: either ADHD domain | 47 of 87 (54%) | 42 of 79 (53%) |
| Cyclical ADHD by informant-confirmed diagnosis (yes vs no) | 69% vs 36% (Fisher P = .004) | 69% vs 34% (Fisher P = .003) |
| DSM-5 count swing among cyclical, mean (median): Inattention | 2.6 (2.5) | 2.5 (2.2) |
| DSM-5 count swing among cyclical, mean (median): Hyperactivity-impulsivity | 2.1 (1.4) | 2.2 (1.4) |
| Cyclical on ADHD alone (of cyclical ADHD) | 16 of 47 (34%) | 14 of 42 (33%) |
| Cyclical in no domain | 25 of 87 (29%) | 23 of 79 (29%) |

Restricted = participants with informant-confirmed ADHD or at least 5 SCID-5 symptoms in either domain (79 of 87); excluded are the 8 whose DSM-5 status rested on daily ratings. Population models were not refit. The 37 who met the SCID-5 criterion without informant confirmation had the same severity as the 42 confirmed participants (median SCID-5 total, 14 vs 13.5; median daily inattention, 1.08 vs 1.08).

### eTable 3: Within-person reliability of the daily composites

| Composite | ω-total (within) |
|---|---|
| Inattention | 0.84 |
| Hyperactivity-impulsivity | 0.81 |
| Negative affect | 0.91 |

Multilevel ω from within-person factor analyses of the daily items (113 participants; 5,583 person-days). ω-hierarchical ranged from 0.59 to 0.74 within person and from 0.80 to 0.90 between persons. In a joint factor analysis of the 18 ADHD items and 21 DRSP items, ADHD and affect items loaded on separate factors (correlations, 0.2 to 0.4); only DRSP difficulty concentrating loaded with inattention.

### eTable 4: Daily hormones and symptoms

#### (A) Each hormone alone: within-person 5-day rolling level.

| Outcome | Hormone | Days / n | b (within) | P | P (BH) | ICC |
|---|---|---|---|---|---|---|
| Inattention | E2 | 2347 / 96 | −0.025 | .07 | .16 | 0.57 |
| Hyperactivity-impulsivity | E2 | 2347 / 96 | −0.028 | .03 | .09 | 0.62 |
| Negative affect | E2 | 2350 / 96 | −0.017 | .28 | .38 | 0.46 |
| Joint/muscle pain | E2 | 2341 / 96 | −0.040 | .03 | .09 | 0.28 |
| Inattention | P4 | 2357 / 96 | −0.021 | .11 | .37 | 0.58 |
| Hyperactivity-impulsivity | P4 | 2357 / 96 | −0.031 | .01 | .11 | 0.62 |
| Negative affect | P4 | 2360 / 96 | −0.003 | .85 | .96 | 0.46 |
| Joint/muscle pain | P4 | 2351 / 96 | −0.027 | .12 | .37 | 0.29 |
| Inattention, robustly cyclical | E2 | 964 / 37 | −0.039 | .07 | .34 | 0.52 |
| Hyperactivity-impulsivity, robustly cyclical | E2 | 588 / 23 | −0.077 | .004 | .03 | 0.58 |
| Inattention, robustly cyclical | P4 | 980 / 37 | −0.021 | .30 | .63 | 0.55 |
| Hyperactivity-impulsivity, robustly cyclical | P4 | 588 / 23 | −0.086 | <.001 | .003 | 0.58 |

Outcomes standardized. b = coefficient for the within-person 5-day rolling hormone, with the person mean as a covariate and a person random intercept; ICC = share of outcome variance between persons; BH within each hormone. Robustly cyclical rows refit the model in participants robustly cyclical on that outcome.

#### (B) Estradiol, progesterone and their product (person-standardized hormones).

| Hormones | Outcome | Term | People / days | β (log scale, per SD) | Ratio per SD | P | q (BH, 4 outcomes) |
|---|---|---|---|---|---|---|---|
| same day | Inattention | E2 | 97 / 2228 | −0.0054 | 0.995 | .65 | .86 |
| same day | Inattention | P4 | 97 / 2228 | −0.0137 | 0.986 | .23 | .59 |
| same day | Inattention | E2 × P4 | 97 / 2228 | 0.0176 | 1.018 | .07 | .26 |
| same day | Hyperactivity-impulsivity | E2 | 97 / 2228 | −0.0193 | 0.981 | .21 | .42 |
| same day | Hyperactivity-impulsivity | P4 | 97 / 2228 | −0.0067 | 0.993 | .65 | .86 |
| same day | Hyperactivity-impulsivity | E2 × P4 | 97 / 2228 | −0.0032 | 0.997 | .78 | .78 |
| same day | Negative affect | E2 | 97 / 2230 | −0.0003 | 1.000 | .98 | .98 |
| same day | Negative affect | P4 | 97 / 2230 | −0.0014 | 0.999 | .89 | .89 |
| same day | Negative affect | E2 × P4 | 97 / 2230 | 0.0111 | 1.011 | .13 | .26 |
| same day | Joint/muscle pain | E2 | 97 / 2220 | −0.0235 | 0.977 | .13 | .42 |
| same day | Joint/muscle pain | P4 | 97 / 2220 | −0.0166 | 0.984 | .29 | .59 |
| same day | Joint/muscle pain | E2 × P4 | 97 / 2220 | 0.0031 | 1.003 | .77 | .78 |
| 5-day rolling | Inattention | E2 | 96 / 2338 | −0.0076 | 0.992 | .62 | .62 |
| 5-day rolling | Inattention | P4 | 96 / 2338 | −0.0187 | 0.981 | .17 | .31 |
| 5-day rolling | Inattention | E2 × P4 | 96 / 2338 | 0.0185 | 1.019 | .14 | .19 |
| 5-day rolling | Hyperactivity-impulsivity | E2 | 96 / 2338 | −0.0117 | 0.988 | .51 | .62 |
| 5-day rolling | Hyperactivity-impulsivity | P4 | 96 / 2338 | −0.0202 | 0.980 | .23 | .31 |
| 5-day rolling | Hyperactivity-impulsivity | E2 × P4 | 96 / 2338 | 0.0133 | 1.013 | .36 | .36 |
| 5-day rolling | Negative affect | E2 | 96 / 2341 | −0.0171 | 0.983 | .09 | .35 |
| 5-day rolling | Negative affect | P4 | 96 / 2341 | 0.0034 | 1.003 | .74 | .74 |
| 5-day rolling | Negative affect | E2 × P4 | 96 / 2341 | 0.0158 | 1.016 | .07 | .18 |
| 5-day rolling | Joint/muscle pain | E2 | 96 / 2332 | −0.0144 | 0.986 | .46 | .62 |
| 5-day rolling | Joint/muscle pain | P4 | 96 / 2332 | −0.0334 | 0.967 | .05 | .22 |
| 5-day rolling | Joint/muscle pain | E2 × P4 | 96 / 2332 | 0.0216 | 1.022 | .09 | .18 |

Mixed model per outcome (Tweedie for the ADHD severities, gamma for negative affect and pain; log link) with a random intercept and random E2 and P4 slopes; BH across the 4 outcomes. For hyperactivity-impulsivity on 5-day rolling hormones, the Tweedie power was fixed at its same-day estimate (1.115).

#### (C) Estradiol-by-progesterone tensor-product surface.

| Hormones | Outcome | People / days | Published form (log1p, Gaussian): edf, P (q) | Matched form (Tweedie/gamma + random slopes): edf, P (q) |
|---|---|---|---|---|
| same day | Inattention | 97 / 2228 | 1, .02 (.09) | 1, .07 (.19) |
| same day | Hyperactivity-impulsivity | 97 / 2228 | 1, .56 (.72) | 5.41, .09 (.19) |
| same day | Negative affect | 97 / 2230 | 4.08, .22 (.43) | 4.21, .18 (.24) |
| same day | Joint/muscle pain | 97 / 2220 | 2.6, .72 (.72) | 3.8, .50 (.50) |
| 5-day rolling | Inattention | 96 / 2338 | 2.27, .08 (.08) | 1.86, .15 (.20) |
| 5-day rolling | Hyperactivity-impulsivity | 96 / 2338 | 2.21, .02 (.049) | 1.11, .60 (.60) |
| 5-day rolling | Negative affect | 96 / 2341 | 1, .004 (.02) | 1, .005 (.01) |
| 5-day rolling | Joint/muscle pain | 96 / 2332 | 4.99, .06 (.08) | 2.75, .006 (.01) |

P for the ti(E2, P4) term, with both main effects in the model. Published form: log(y + 1), Gaussian, random intercept. Matched form: the families and random slopes of panel B.

#### (D) Daily hormone change.

| Outcome | Hormone change | Day | Random intercept and change slopes (primary), β (P) | Random intercept only, P |
|---|---|---|---|---|
| Inattention | E2 change | previous day | +0.009 (.38) | .38 |
| Inattention | E2 change | same day | +0.008 (.44) | .35 |
| Inattention | P4 change | previous day | −0.020 (.11) | .04 |
| Inattention | P4 change | same day | −0.005 (.75) | .24 |
| Hyperactivity-impulsivity | E2 change | previous day | +0.025 (.14) | .08 |
| Hyperactivity-impulsivity | E2 change | same day | −0.011 (.43) | .21 |
| Hyperactivity-impulsivity | P4 change | previous day | −0.006 (.78) | .24 |
| Hyperactivity-impulsivity | P4 change | same day | −0.012 (.55) | .63 |
| Negative affect | E2 change | previous day | −0.010 (.23) | .19 |
| Negative affect | E2 change | same day | −0.013 (.17) | .08 |
| Negative affect | P4 change | previous day | −0.024 (.01) | .003 |
| Negative affect | P4 change | same day | −0.013 (.14) | .11 |
| Joint/muscle pain | E2 change | previous day | −0.005 (.69) | .63 |
| Joint/muscle pain | E2 change | same day | −0.023 (.06) | .06 |
| Joint/muscle pain | P4 change | previous day | −0.019 (.15) | .12 |
| Joint/muscle pain | P4 change | same day | −0.013 (.35) | .21 |

Change = daily change in the participant's 5-day rolling hormone, person-standardized; negative β = worse as the hormone falls. Families as in Table 2. Cycle time is not in these models (eMethods 5).

### eTable 5: Co-occurrence of robust cyclicity across domains

| Domain A | Domain B | Both | A only | B only | Neither | φ |
|---|---|---|---|---|---|---|
| Inattention | Hyperactivity-impulsivity | 13 | 24 | 10 | 40 | 0.17 |
| Inattention | Negative affect | 23 | 14 | 6 | 44 | 0.53 |
| Inattention | Pain | 12 | 25 | 17 | 33 | −0.02 |
| Hyperactivity-impulsivity | Negative affect | 11 | 12 | 18 | 46 | 0.18 |
| Hyperactivity-impulsivity | Pain | 13 | 10 | 16 | 48 | 0.29 |
| Negative affect | Pain | 12 | 17 | 17 | 41 | 0.12 |

φ = correlation between the 2 cyclicity flags. Of the 47 cyclical on either ADHD domain, 31 (66%) were also cyclical on negative affect or pain; 25 of 87 were cyclical on no domain and 11 on all 3.

### eTable 6: Robust cyclicity by informant-confirmed ADHD

#### (A) Robust cyclicity by informant-confirmed diagnosis.

| Robust cyclicity on | Informant-confirmed ADHD (n = 42) | Not confirmed (n = 39) | OR (95% CI) | P | P (BH, 5 contrasts) |
|---|---|---|---|---|---|
| Inattention | 23/42 (55%) | 10/39 (26%) | 3.45 (1.25-10.12) | .01 | .03 |
| Hyperactivity-impulsivity | 15/42 (36%) | 7/39 (18%) | 2.51 (0.82-8.41) | .09 | .14 |
| Negative affect | 16/42 (38%) | 10/39 (26%) | 1.77 (0.63-5.21) | .25 | .31 |
| Joint/muscle pain | 15/42 (36%) | 12/39 (31%) | 1.25 (0.45-3.52) | .81 | .81 |
| Either ADHD domain | 29/42 (69%) | 14/39 (36%) | 3.91 (1.44-11.17) | .004 | .02 |

**(B) Still robustly cyclical after within-person adjustment for same-day negative affect, by confirmation.**

| Domain (robustly cyclical participants) | Still robust after same-day affect adjustment: informant-confirmed | Not confirmed | OR (95% CI) | P | P (BH, 2 domains) | Median % of cyclic $R^2$ retained: confirmed / not |
|---|---|---|---|---|---|---|
| Inattention | 15/23 (65%) | 3/10 (30%) | 4.17 (0.71-32.14) | .13 | .25 | 79 / 19 |
| Hyperactivity-impulsivity | 9/15 (60%) | 5/7 (71%) | 0.61 (0.04-5.53) | >.99 | >.99 | 75 / 76 |

(A) Participants with informant data (81 of 87); Fisher exact tests; BH across 5 contrasts. (B) Robustly cyclical participants; still robust = still significant against the participant's own permutation null after adjustment; BH across 2 domains. Participants whose unadjusted refit did not reproduce their cyclicity are counted as retaining 0% (eTable 10).

### eTable 7: Participant characteristics and cyclicity as a continuous exposure

| Family | Characteristic | n | Metric | Estimate | P | P (BH within family) | Binary-flag version |
|---|---|---|---|---|---|---|---|
| C-PASS | C-PASS MRMD (any evaluable cycle) | 77 | amplitude | OR per SD 1.05 (0.52-2.10) | .89 | .89 | 9% vs 12%; OR 0.73; P = .72 |
| C-PASS | C-PASS MRMD (any evaluable cycle) | 77 | cyclic $R^2$ | OR per SD 0.88 (0.41-1.89) | .75 | >.99 | 9% vs 12%; OR 0.73; P = .72 |
| C-PASS | C-PASS PME, MRMD or PMDD (any evaluable cycle) | 77 | amplitude | OR per SD 1.20 (0.65-2.19) | .56 | .89 | 11% vs 12%; OR 0.93; P > .99 |
| C-PASS | C-PASS PME, MRMD or PMDD (any evaluable cycle) | 77 | cyclic $R^2$ | OR per SD 1.00 (0.49-2.04) | >.99 | >.99 | 11% vs 12%; OR 0.93; P > .99 |
| C-PASS | C-PASS core-item premenstrual change (%) | 77 | amplitude | Spearman ρ 0.02 | .85 | .89 | median 1.1 vs 3.3; Wilcoxon P = .46 |
| C-PASS | C-PASS core-item premenstrual change (%) | 77 | cyclic $R^2$ | Spearman ρ −0.05 | .67 | >.99 | median 1.1 vs 3.3; Wilcoxon P = .46 |
| DSM-5 count swing | Hyperactivity-impulsivity count swing (symptoms) | 87 | amplitude | +0.52 symptoms per 0.10 amplitude (r = 0.79) | <.001 | <.001 | median 1.4 (cyclical, n = 23) vs 0.2 |
| DSM-5 count swing | Hyperactivity-impulsivity count swing (symptoms) | 87 | cyclic $R^2$ | +0.36 symptoms per 0.10 cyclic $R^2$ (r = 0.62) | <.001 | <.001 | median 1.4 (cyclical, n = 23) vs 0.2 |
| DSM-5 count swing | Inattention count swing (symptoms) | 87 | amplitude | +0.59 symptoms per 0.10 amplitude (r = 0.79) | <.001 | <.001 | median 2.5 (cyclical, n = 37) vs 0.0 |
| DSM-5 count swing | Inattention count swing (symptoms) | 87 | cyclic $R^2$ | +0.47 symptoms per 0.10 cyclic $R^2$ (r = 0.70) | <.001 | <.001 | median 2.5 (cyclical, n = 37) vs 0.0 |
| Demographic / data | Age | 83 | amplitude | Spearman ρ −0.07 | .55 | .75 | median 21.1 vs 21.4; Wilcoxon P > .99 |

| Family | Characteristic | n | Metric | Estimate | P | P (BH within family) | Binary-flag version |
|---|---|---|---|---|---|---|---|
| Demographic / data | Age | 83 | cyclic $R^2$ | Spearman ρ −0.04 | .72 | .72 | median 21.1 vs 21.4; Wilcoxon P > .99 |
| Demographic / data | Body mass index | 82 | amplitude | Spearman ρ 0.04 | .75 | .75 | median 24.3 vs 25.0; Wilcoxon P = .65 |
| Demographic / data | Body mass index | 82 | cyclic $R^2$ | Spearman ρ 0.11 | .33 | .44 | median 24.3 vs 25.0; Wilcoxon P = .65 |
| Demographic / data | Daily-survey completion (%) | 87 | amplitude | Spearman ρ −0.05 | .64 | .75 | median 66.7 vs 66.6; Wilcoxon P = .58 |
| Demographic / data | Daily-survey completion (%) | 87 | cyclic $R^2$ | Spearman ρ −0.17 | .11 | .21 | median 66.7 vs 66.6; Wilcoxon P = .58 |
| Demographic / data | Days with hormone sample | 87 | amplitude | Spearman ρ −0.19 | .07 | .30 | median 30.0 vs 32.5; Wilcoxon P = .22 |
| Demographic / data | Days with hormone sample | 87 | cyclic $R^2$ | Spearman ρ −0.34 | .001 | .006 | median 30.0 vs 32.5; Wilcoxon P = .22 |
| Diagnosis | Any anxiety disorder | 78 | amplitude | OR per SD 1.04 (0.68-1.60) | .86 | .86 | 53% vs 53%; OR 0.99; P > .99 |
| Diagnosis | Any anxiety disorder | 78 | cyclic $R^2$ | OR per SD 0.89 (0.57-1.38) | .59 | .78 | 53% vs 53%; OR 0.99; P > .99 |
| Diagnosis | Any depressive disorder | 78 | amplitude | OR per SD 0.57 (0.28-1.17) | .13 | .46 | 18% vs 26%; OR 0.60; P = .42 |
| Diagnosis | Any depressive disorder | 78 | cyclic $R^2$ | OR per SD 0.76 (0.44-1.34) | .35 | .76 | 18% vs 26%; OR 0.60; P = .42 |
| Diagnosis | Generalized anxiety disorder | 78 | amplitude | OR per SD 1.30 (0.82-2.07) | .27 | .69 | 33% vs 21%; OR 1.79; P = .31 |
| Diagnosis | Generalized anxiety disorder | 78 | cyclic $R^2$ | OR per SD 1.48 (0.90-2.44) | .13 | .46 | 33% vs 21%; OR 1.79; P = .31 |
| Diagnosis | Informant-confirmed ADHD diagnosis | 81 | amplitude | OR per SD 1.75 (1.02-3.00) | .04 | .46 | 67% vs 34%; OR 3.91; P = .004 |
| Diagnosis | Informant-confirmed ADHD diagnosis | 81 | cyclic $R^2$ | OR per SD 1.90 (1.17-3.09) | .01 | .11 | 67% vs 34%; OR 3.91; P = .004 |
| Diagnosis | PTSD | 78 | amplitude | OR per SD 1.50 (0.93-2.45) | .10 | .46 | 33% vs 26%; OR 1.34; P = .62 |
| Diagnosis | PTSD | 78 | cyclic $R^2$ | OR per SD 1.53 (0.93-2.49) | .09 | .46 | 33% vs 26%; OR 1.34; P = .62 |

| Family | Characteristic | n | Metric | Estimate | P | P (BH within family) | Binary-flag version |
|---|---|---|---|---|---|---|---|
| Diagnosis | Provisional SCID PMDD | 78 | amplitude | OR per SD 1.26 (0.80-1.98) | .33 | .69 | 55% vs 45%; OR 1.50; P = .50 |
| Diagnosis | Provisional SCID PMDD | 78 | cyclic $R^2$ | OR per SD 0.92 (0.59-1.43) | .71 | .78 | 55% vs 45%; OR 1.50; P = .50 |
| Diagnosis | SCID hyperactivity-impulsivity count | 78 | amplitude | Spearman ρ 0.08 | .48 | .69 | median 5.0 vs 5.0; Wilcoxon P = .69 |
| Diagnosis | SCID hyperactivity-impulsivity count | 78 | cyclic $R^2$ | Spearman ρ −0.06 | .59 | .78 | median 5.0 vs 5.0; Wilcoxon P = .69 |
| Diagnosis | SCID inattention count | 78 | amplitude | Spearman ρ 0.05 | .66 | .73 | median 8.0 vs 8.0; Wilcoxon P > .99 |
| Diagnosis | SCID inattention count | 78 | cyclic $R^2$ | Spearman ρ −0.02 | .85 | .85 | median 8.0 vs 8.0; Wilcoxon P > .99 |
| Diagnosis | SCID total count | 78 | amplitude | Spearman ρ 0.08 | .48 | .69 | median 13.0 vs 13.0; Wilcoxon P = .79 |
| Diagnosis | SCID total count | 78 | cyclic $R^2$ | Spearman ρ −0.05 | .65 | .78 | median 13.0 vs 13.0; Wilcoxon P = .79 |
| Diagnosis | Social anxiety disorder | 78 | amplitude | OR per SD 1.17 (0.74-1.86) | .50 | .69 | 33% vs 21%; OR 1.79; P = .31 |
| Diagnosis | Social anxiety disorder | 78 | cyclic $R^2$ | OR per SD 1.21 (0.74-1.98) | .45 | .78 | 33% vs 21%; OR 1.79; P = .31 |
| Diagnosis | Substance use disorder | 78 | amplitude | OR per SD 1.15 (0.68-1.95) | .60 | .73 | 18% vs 16%; OR 1.13; P > .99 |
| Diagnosis | Substance use disorder | 78 | cyclic $R^2$ | OR per SD 1.36 (0.76-2.43) | .30 | .76 | 18% vs 16%; OR 1.13; P > .99 |
| Medication | Antidepressant (any) | 78 | amplitude | OR per SD 1.42 (0.88-2.29) | .15 | .22 | 35% vs 18%; OR 2.36; P = .13 |
| Medication | Antidepressant (any) | 78 | cyclic $R^2$ | OR per SD 1.45 (0.88-2.39) | .14 | .21 | 35% vs 18%; OR 2.36; P = .13 |
| Medication | SSRI | 78 | amplitude | OR per SD 1.69 (0.99-2.88) | .06 | .17 | 28% vs 11%; OR 3.18; P = .08 |
| Medication | SSRI | 78 | cyclic $R^2$ | OR per SD 1.65 (0.94-2.91) | .08 | .21 | 28% vs 11%; OR 3.18; P = .08 |
| Medication | Stimulant use | 80 | amplitude | OR per SD 1.23 (0.75-2.01) | .42 | .42 | 24% vs 16%; OR 1.66; P = .41 |
| Medication | Stimulant use | 80 | cyclic $R^2$ | OR per SD 1.15 (0.67-1.98) | .62 | .62 | 24% vs 16%; OR 1.66; P = .41 |

### eTable 8: Cognitive performance aligned with each participant's symptom cycle

| Outcome | Symptom cycle | n / visits | Within-person β per own-SD (SE) | P | P (BH, 4 composite pairings) | Between-person β (P) | Unconstrained matched terms, cos and sin (joint P) | Unshrunk per-person coefficients: β (P) |
|---|---|---|---|---|---|---|---|---|
| Working memory (composite) | own inattention cycle | 85 / 321 | 0.27 (0.13) | .03 | .13 | 2.11 (.03) | 0.26, 0.25 (.13) | 0.10 (.13) |
| Working memory (composite) | own hyperactivity-impulsivity cycle | 85 / 321 | 0.07 (0.15) | .65 | .68 | 1.77 (.15) | 0.14, −0.10 (.71) | 0.05 (.47) |
| Inhibition (composite) | own inattention cycle | 85 / 314 | −0.06 (0.15) | .68 | .68 | −0.30 (.65) | −0.27, 0.25 (.26) | −0.04 (.60) |
| Inhibition (composite) | own hyperactivity-impulsivity cycle | 85 / 314 | −0.15 (0.17) | .39 | .68 | −0.39 (.63) | −0.20, −0.06 (.61) | −0.11 (.16) |
| N-back | own inattention cycle | 85 / 320 | 0.19 (0.16) | .22 | NA | 0.86 (.37) | 0.22, 0.13 (.50) | 0.06 (.49) |
| Symmetry span | own inattention cycle | 85 / 311 | 0.20 (0.15) | .21 | NA | 2.22 (.02) | 0.07, 0.35 (.33) | 0.10 (.25) |
| Letter-number sequencing | own inattention cycle | 85 / 320 | 0.15 (0.16) | .36 | NA | 1.94 (.04) | 0.18, 0.10 (.64) | 0.05 (.52) |

All scores are deficits (higher = worse). Within-person β = change in the outcome (SD) per own-SD of the participant's predicted symptom at the visit, adjusting for the population cycle, practice, and a person random intercept. Between-person β uses the participant's mean predicted symptom across visits. Unconstrained = the 4 cosine and sine products in place of the single alignment term. Unshrunk = per-person least-squares coefficients. BH across the 4 composite pairings (NA for single tasks).

### eTable 9: Next-day cross-lagged associations of ADHD symptoms and negative affect

| Direction | Participants | Consecutive-day pairs | β (95% CI) | P | P (BH) |
|---|---|---|---|---|---|
| Negative affect → Inattention | 109 | 2,235 | 0.046 (0.004-0.088) | .03 | .03 |
| Inattention → Negative affect | 109 | 2,235 | 0.054 (0.012-0.097) | .01 | .02 |
| Negative affect → Hyperactivity-impulsivity | 109 | 2,235 | 0.071 (0.031-0.110) | <.001 | .002 |
| Hyperactivity-impulsivity → Negative affect | 109 | 2,235 | 0.067 (0.026-0.108) | .001 | .003 |

Linear mixed model of the outcome on its own previous-day value and the other domain's previous-day value, with a person random intercept; variables person-centered and scaled to the within-person SD. Consecutive-day pairs only. Wald 95% CI; BH across the 4 directions.

### eTable 10: ADHD cyclicity after adjustment for negative affect or pain

| Outcome | Covariate removed within person | Robustly cyclical on record (evaluable) | Still robustly cyclical after adjustment | Median % of cyclic $R^2$ retained (IQR) | Kept ≥ 50% | Median cyclic $R^2$, % (before → after) |
|---|---|---|---|---|---|---|
| Inattention | same-day negative affect | 37 | 22 (59%) | 79 (20-101) | 23 | 52 → 28 |
| Inattention | previous-day negative affect | 35 | 15 (43%) | 78 (15-92) | 23 | 46 → 30 |
| Inattention | same-day pain | 37 | 31 (84%) | 98 (79-100) | 32 | 52 → 42 |
| Hyperactivity-impulsivity | same-day negative affect | 23 | 15 (65%) | 76 (48-100) | 17 | 44 → 32 |
| Hyperactivity-impulsivity | previous-day negative affect | 22 | 11 (50%) | 67 (19-96) | 13 | 42 → 29 |
| Hyperactivity-impulsivity | same-day pain | 23 | 16 (70%) | 94 (68-101) | 18 | 44 → 37 |

For each robustly cyclical participant, the curve was refit after same-day negative affect, previous-day negative affect, or same-day pain was removed within person, and tested against a within-person permutation null (2,000 shuffles). Share retained = adjusted over unadjusted cyclic $R^2$ on the same days. The unadjusted refit reproduced 34 of 37 inattention and 22 of 23 hyperactivity-impulsivity classifications; the others are counted as retaining 0%. Restricted to reproduced participants, 21 of 34 (62%) and 15 of 22 (68%) remained robust after same-day affect adjustment. At the population level, adjusting for same-day negative affect reduced the inattention cycle to 30% of its amplitude and the hyperactivity-impulsivity cycle to 88%. Because affect also tracks the cycle, these adjustments cannot separate confounding from a shared cause.

### eTable 11: Prospective PMDD (C-PASS)

| Measure | Value |
|---|---|
| Participants assessed for cyclicity | 87 |
| Participants with ≥1 scorable cycle | 77 (62 menses-anchored, 15 ovulation-anchored) |
| Scorable cycles | 80 |
| Cycles meeting PMDD criteria | 0 |
| Cycles meeting MRMD criteria | 8 (in 8 participants) |
| Cycles with premenstrual exacerbation (PME) | 3 (in 3 participants) |
| Participants meeting the two-cycle rule for PMDD or MRMD | 0 |

Premenstrual days −7 to −1 vs postmenstrual days 4 to 10; elevation of at least 30% of the item's scale range; each participant scored with the anchoring (menses or ovulation) that gave more scorable cycles. eTable 7 counts participants with any category (n = 9). MRMD indicates menstrually related mood disorder; PME, premenstrual exacerbation; PMDD, premenstrual dysphoric disorder.

### eTable 12: Retrospective PMDD report: items, and agreement with daily ratings by peak window

#### (A) Individual retrospective SCID-5 PMDD items and robust ADHD cyclicity.

| Robustly cyclical on | SCID PMDD item | Endorsers / non-endorsers | % cyclical (endorsers / non) | OR (95% CI) | P | P (BH, 11 items) |
|---|---|---|---|---|---|---|
| Inattention | Sleep changes (hypersomnia/insomnia) | 37 / 35 | 51% / 23% | 3.50 (1.16-11.39) | .02 | .17 |
| Inattention | Decreased interest (anhedonia) | 44 / 24 | 43% / 25% | 2.25 (0.68-8.33) | .19 | .64 |
| Inattention | Fatigue / low energy | 61 / 16 | 39% / 31% | 1.42 (0.39-5.89) | .77 | .99 |
| Inattention | Irritability / anger | 54 / 18 | 39% / 33% | 1.27 (0.37-4.78) | .78 | .99 |
| Inattention | Physical symptoms | 60 / 14 | 38% / 36% | 1.12 (0.29-4.80) | >.99 | >.99 |
| Inattention | Difficulty concentrating | 51 / 25 | 37% / 36% | 1.06 (0.35-3.28) | >.99 | >.99 |
| Inattention | Affective lability | 57 / 20 | 37% / 40% | 0.88 (0.28-2.90) | .80 | .99 |
| Inattention | Appetite change / cravings | 34 / 36 | 35% / 39% | 0.86 (0.29-2.52) | .81 | .99 |
| Inattention | Anxiety / tension | 38 / 33 | 32% / 39% | 0.71 (0.24-2.11) | .62 | .99 |
| Inattention | Overwhelmed / out of control | 38 / 35 | 29% / 43% | 0.55 (0.18-1.59) | .23 | .64 |
| Inattention | Depressed mood | 50 / 26 | 30% / 50% | 0.43 (0.14-1.28) | .13 | .64 |
| Hyperactivity-impulsivity | Irritability / anger | 54 / 18 | 31% / 11% | 3.62 (0.72-36.00) | .13 | .97 |
| Hyperactivity-impulsivity | Difficulty concentrating | 51 / 25 | 29% / 16% | 2.17 (0.58-10.16) | .27 | .97 |
| Hyperactivity-impulsivity | Anxiety / tension | 38 / 33 | 29% / 21% | 1.50 (0.45-5.34) | .59 | .97 |
| Hyperactivity-impulsivity | Appetite change / cravings | 34 / 36 | 29% / 25% | 1.25 (0.38-4.12) | .79 | .97 |
| Hyperactivity-impulsivity | Decreased interest (anhedonia) | 44 / 24 | 27% / 25% | 1.12 (0.32-4.30) | >.99 | >.99 |

| Robustly cyclical on | SCID PMDD item | Endorsers / non-endorsers | % cyclical (endorsers / non) | OR (95% CI) | P | P (BH, 11 items) |
|---|---|---|---|---|---|---|
| Hyperactivity-impulsivity | Physical symptoms | 60 / 14 | 27% / 29% | 0.91 (0.22-4.55) | >.99 | >.99 |
| Hyperactivity-impulsivity | Sleep changes (hypersomnia/insomnia) | 37 / 35 | 24% / 29% | 0.81 (0.25-2.61) | .79 | .97 |
| Hyperactivity-impulsivity | Overwhelmed / out of control | 38 / 35 | 24% / 29% | 0.78 (0.24-2.52) | .79 | .97 |
| Hyperactivity-impulsivity | Affective lability | 57 / 20 | 25% / 30% | 0.76 (0.22-2.90) | .77 | .97 |
| Hyperactivity-impulsivity | Fatigue / low energy | 61 / 16 | 25% / 31% | 0.72 (0.19-3.08) | .75 | .97 |
| Hyperactivity-impulsivity | Depressed mood | 50 / 26 | 22% / 31% | 0.64 (0.19-2.17) | .42 | .97 |

Participants with SCID-5 module data (n = 78); BH across the 11 items within each domain. The SCID-5 sleep item combines hypersomnia and insomnia.

#### (B) Retrospective report versus prospective daily ratings, by each participant's peak window.

| Outcome | Group | n | Reported PMDD retrospectively (SCID-5), n (%) | Premenstrual elevation ≥30% on daily ratings (C-PASS), n/N (%) |
|---|---|---|---|---|
| Hyperactivity-impulsivity | Not robustly cyclical | 58 | 28 (48%) | 9/50 (18%) |
| Hyperactivity-impulsivity | Menstrual peak | 9 | 6 (67%) | 1/9 (11%) |
| Hyperactivity-impulsivity | Luteal peak | 5 | 2 (40%) | 4/4 (100%) |
| Hyperactivity-impulsivity | Peri-ovulatory peak | 6 | 3 (50%) | 1/6 (17%) |
| Inattention | Not robustly cyclical | 48 | 22 (46%) | 9/40 (23%) |
| Inattention | Menstrual peak | 15 | 11 (73%) | 1/15 (7%) |
| Inattention | Luteal peak | 8 | 3 (38%) | 5/8 (63%) |
| Inattention | Peri-ovulatory peak | 7 | 3 (43%) | 0/6 (0%) |
| Joint/muscle pain | Not robustly cyclical | 54 | 28 (52%) | 8/48 (17%) |
| Joint/muscle pain | Menstrual peak | 12 | 5 (42%) | 4/10 (40%) |
| Joint/muscle pain | Luteal peak | 6 | 3 (50%) | 1/6 (17%) |
| Joint/muscle pain | Peri-ovulatory peak | 6 | 3 (50%) | 2/5 (40%) |
| Negative affect | Not robustly cyclical | 54 | 22 (41%) | 9/46 (20%) |
| Negative affect | Menstrual peak | 10 | 8 (80%) | 1/10 (10%) |
| Negative affect | Luteal peak | 9 | 5 (56%) | 5/8 (63%) |
| Negative affect | Peri-ovulatory peak | 5 | 4 (80%) | 0/5 (0%) |

Groups: participants not robustly cyclical on the outcome, and robustly cyclical participants by peak window (as in Table 2). Retrospective = provisional PMDD on the SCID-5 at intake. Prospective = premenstrual elevation of at least 30% on at least 1 DRSP item (the C-PASS criterion), among participants with a scorable cycle (N). κ = 0.05 (69 participants with both).

### eTable 13: Medication and cyclicity

| Medication | Outcome | Users / non-users | Days | Cycle difference: edf | P | q (BH, 8 tests) | Level ratio, users vs not (P) |
|---|---|---|---|---|---|---|---|
| SSRI | Inattention | 19 / 74 | 2280 | 0 | .71 | .81 | 1.02 (.82) |
| SSRI | Hyperactivity-impulsivity | 19 / 74 | 2280 | 0 | .59 | .78 | 1.03 (.84) |
| SSRI | Negative affect | 19 / 74 | 2282 | 0 | .26 | .53 | 1.08 (.23) |
| SSRI | Joint/muscle pain | 19 / 74 | 2272 | 0 | .84 | .84 | 0.94 (.41) |
| Stimulant | Inattention | 23 / 69 | 2335 | 1.67 | .004 | .02 | 0.86 (.16) |
| Stimulant | Hyperactivity-impulsivity | 23 / 69 | 2335 | 3.67 | .003 | .02 | 0.81 (.09) |
| Stimulant | Negative affect | 23 / 69 | 2337 | 2.86 | .02 | .046 | 0.97 (.57) |
| Stimulant | Joint/muscle pain | 23 / 69 | 2325 | 0 | .42 | .67 | 0.95 (.40) |

| Medication | Outcome | Users / non-users | Days | Cycle difference: edf | P | q (BH, 8 tests) | Level ratio, users vs not (P) |
|---|---|---|---|---|---|---|---|

| Robustly cyclical on | All | Without SSRI users | SSRI users | P | Without stimulant users | Stimulant users | P | Neither medication |
|---|---|---|---|---|---|---|---|---|
| Inattention | 37 of 87 (43%) | 22 of 63 (35%) | 8 of 15 (53%) | .24 | 23 of 64 (36%) | 10 of 16 (63%) | .09 | 15 of 52 (29%) |
| Hyperactivity-impulsivity | 23 of 87 (26%) | 14 of 63 (22%) | 6 of 15 (40%) | .19 | 18 of 64 (28%) | 3 of 16 (19%) | .54 | 12 of 52 (23%) |
| Negative affect | 29 of 87 (33%) | 18 of 63 (29%) | 6 of 15 (40%) | .53 | 18 of 64 (28%) | 8 of 16 (50%) | .14 | 13 of 52 (25%) |
| Joint/muscle pain | 29 of 87 (33%) | 19 of 63 (30%) | 5 of 15 (33%) | >.99 | 23 of 64 (36%) | 3 of 16 (19%) | .24 | 19 of 52 (37%) |
| Either ADHD domain | 47 of 87 (54%) | 29 of 63 (46%) | 11 of 15 (73%) | .08 | 32 of 64 (50%) | 10 of 16 (63%) | .41 | 22 of 52 (42%) |

Population models: the Table 2 model with a medication-by-cycle difference smooth. Level ratio = users' mean relative to non-users'. Person level: denominators are participants with a medication record.

### eTable 14: Diary nonresponse and missing-data sensitivity

#### (A) Skipping the next day's survey, by the previous day's within-person symptom level.

| Outcome | Person-days | n | β (prior-day within-person level → next-day skip) | OR per SD | z | P |
|---|---|---|---|---|---|---|
| Inattention severity | 2558 | 107 | −0.13 | 0.88 | −2.39 | .02 |
| Hyperactivity-impulsivity severity | 2558 | 107 | −0.04 | 0.97 | −0.67 | .51 |
| Negative affect | 2561 | 107 | −0.09 | 0.91 | −1.65 | .10 |
| Joint/muscle pain | 2549 | 107 | −0.10 | 0.91 | −1.79 | .07 |

Mixed logistic model of next-day skipping; a negative β means fewer skipped days after a higher-symptom day.

#### (B) Population cycle amplitude if missed days differed from what the observed days predict.

| Outcome | Cycle amplitude, observed days only | Missed days assumed 1 SD lower | Change | Missed days assumed 1 SD higher | Change |
|---|---|---|---|---|---|
| Inattention severity | 0.37 | 0.32 | −15% | 0.58 | +55% |
| Hyperactivity-impulsivity severity | 0.23 | 0.20 | −11% | 0.35 | +55% |
| Negative affect | 2.38 | 1.98 | −17% | 3.53 | +48% |
| Joint/muscle pain | 2.33 | 1.31 | −44% | 5.52 | +137% |

Missed days were imputed from a population model of the observed days, shifted 1 within-person SD lower or higher, and the cycle was refit. Amplitude = trough to peak on each outcome's scale. The imputation model is simpler than the Table 2 model, so these amplitudes are not comparable with Table 2.

### eFigures

### eFigure 1: Participant flow

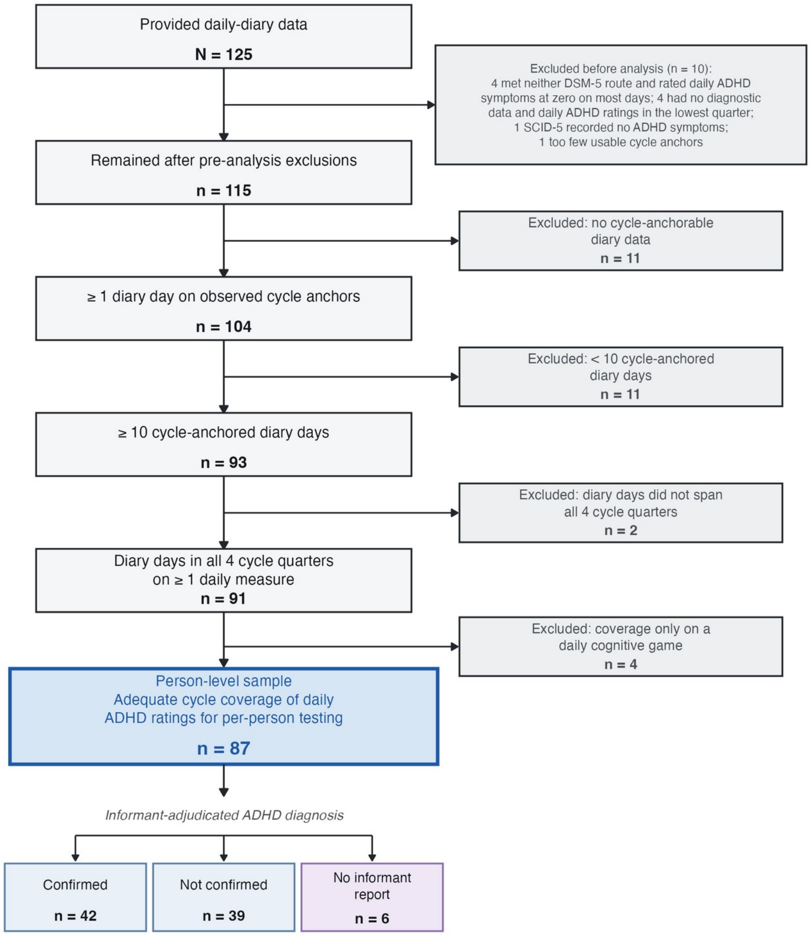


The analytic sample is the 106 with any rated day with cycle time; the person-level sample is the 87 with adequate coverage of the daily ADHD ratings (eMethods 1; eTable 1).

### eFigure 2: Working memory aligned with each participant's inattention cycle

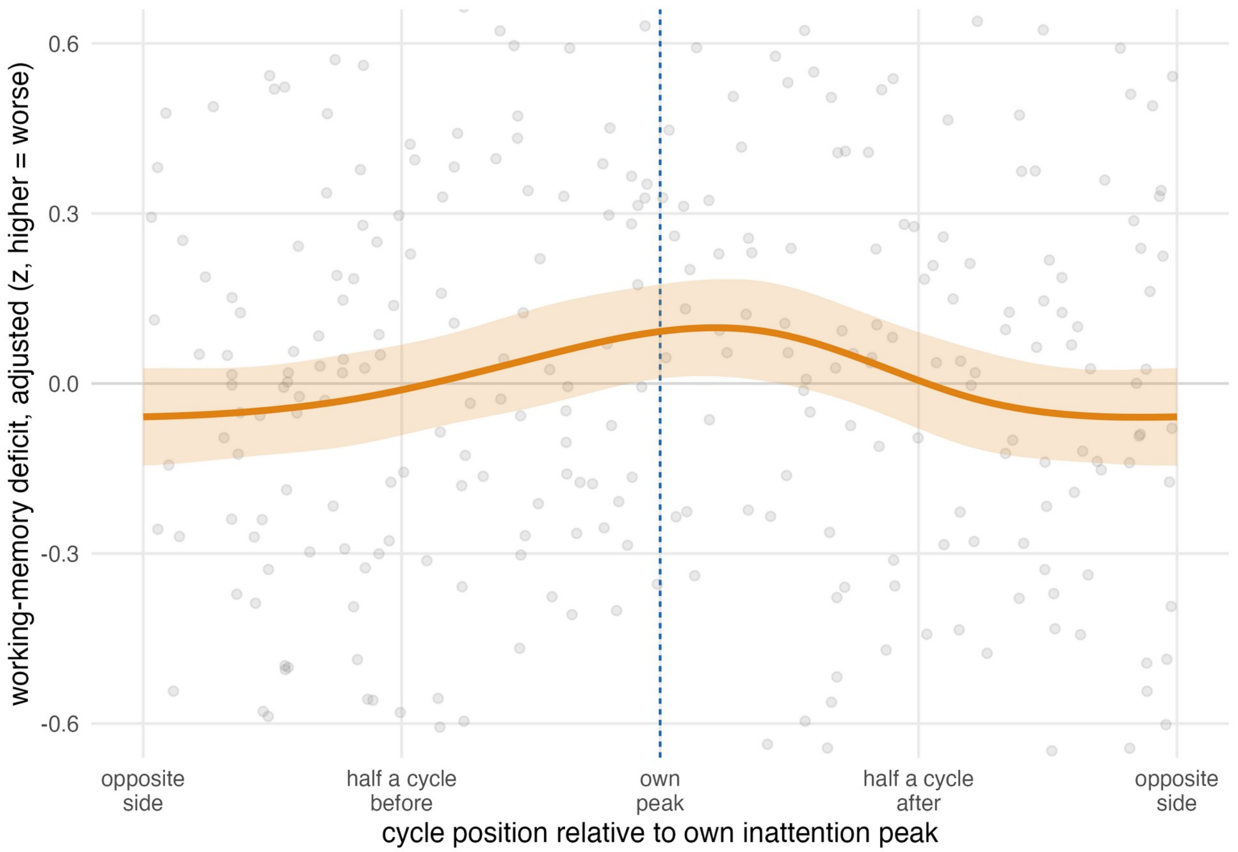


Each lab visit is placed by its distance from the participant's own inattention peak (0 = peak). Gray points = visits (working memory adjusted for practice, own level, and the population cycle; higher = worse); orange line = cyclic smooth across participants (95% CI), weighted by cycle amplitude. Within-person β = 0.27 per own-SD; P = .03 (85 participants, 321 visits; eTable 8).

### eFigure 3: Sample-average cycle by stimulant and SSRI use

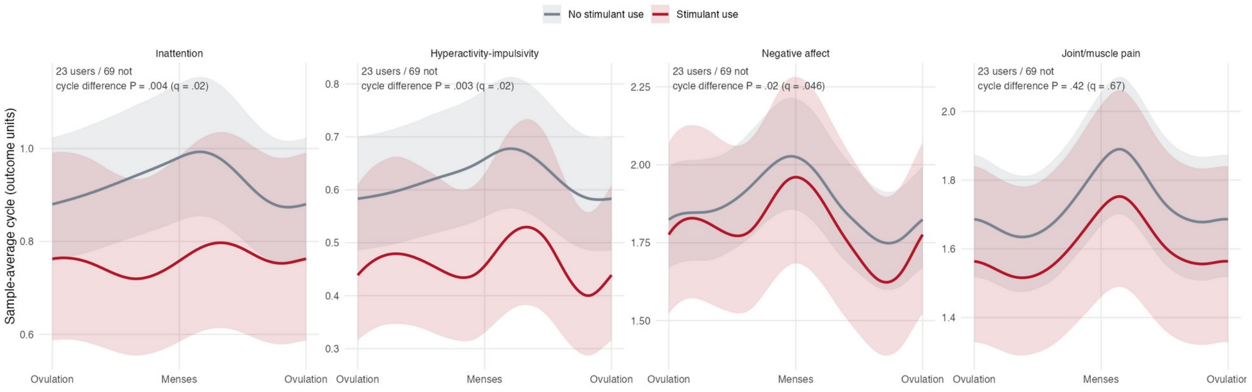


(A) Stimulant use (23 users, 69 non-users; bands = 95% CI). The cycle differed for inattention (P = .004), hyperactivity-impulsivity (P = .003), and negative affect (P = .02), but not pain (eTable 13).

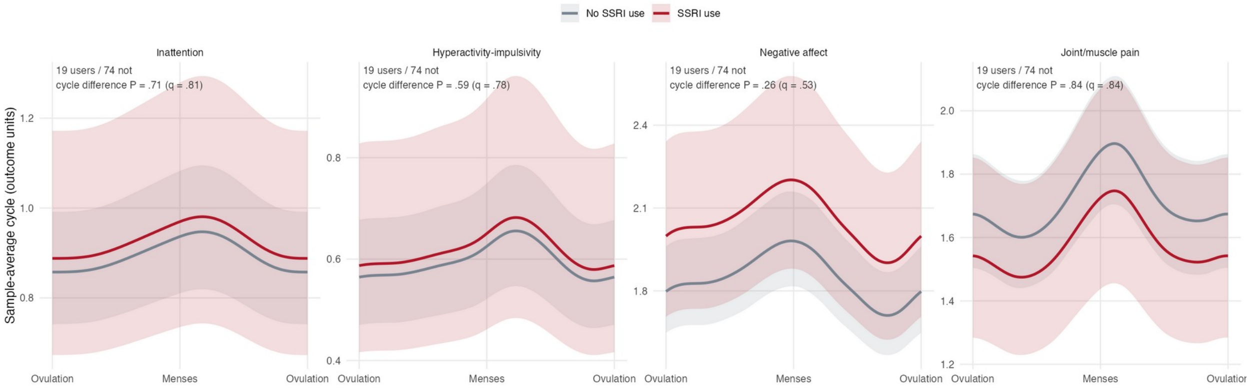


(B) SSRI use (19 users, 74 non-users). The cycle did not differ on any outcome (P = .26 to .84; eTable 13).

### eReferences


1. Nagpal A, Schmalenberger KM, Barone JC, et al. How to study the menstrual cycle as a continuous variable: implementing phase-aligned cycle time scaling (PACTS) with the menstrualcycleR package. Psychoneuroendocrinology. 2025;181:107584. doi:10.1016/j.psyneuen.2025.107584

2. Wood SN. Generalized Additive Models: An Introduction with R. 2nd ed. Chapman & Hall/CRC; 2017.

3. Pedersen EJ, Miller DL, Simpson GL, Ross N. Hierarchical generalized additive models in ecology: an introduction with mgcv. PeerJ. 2019;7:e6876. doi:10.7717/peerj.6876

4. Arslan RC, Blake K, Botzet LJ, et al. Not within spitting distance: salivary immunoassays of estradiol have subpar validity for predicting cycle phase. Psychoneuroendocrinology. 2023;149:105994. doi:10.1016/j.psyneuen.2022.105994

5. Jaeggi SM, Buschkuehl M, Perrig WJ, Meier B. The concurrent validity of the N-back task as a working memory measure. Memory. 2010;18(4):394-412. doi:10.1080/09658211003702171

6. Redick TS, Broadway JM, Meier ME, et al. Measuring working memory capacity with automated complex span tasks. Eur J Psychol Assess. 2012;28(3):164-171. doi:10.1027/1015-5759/a000123

7. Wechsler D. Wechsler Adult Intelligence Scale. 4th ed. Pearson; 2008.

8. Stroop JR. Studies of interference in serial verbal reactions. J Exp Psychol. 1935;18(6):643-662. doi:10.1037/h0054651

9. Hutton SB, Ettinger U. The antisaccade task as a research tool in psychopathology: a critical review. Psychophysiology. 2006;43(3):302-313. doi:10.1111/j.1469-8986.2006.00403.x

10. Wactawski-Wende J, Schisterman EF, Hovey KM, et al. BioCycle study: design of the longitudinal study of the oxidative stress and hormone variation during the menstrual cycle. Paediatr Perinat Epidemiol. 2009;23(2):171-184. doi:10.1111/j.1365-3016.2008.00985.x

11. Eisenlohr-Moul TA, Girdler SS, Schmalenberger KM, et al. Toward the reliable diagnosis of DSM-5 premenstrual dysphoric disorder: the Carolina Premenstrual Assessment Scoring System (C-PASS). Am J Psychiatry. 2017;174(1):51-59. doi:10.1176/appi.ajp.2016.15121510
